# The effect of population selection criteria on model estimates and data missingness in electronic health record studies

**DOI:** 10.1101/2025.02.10.25321999

**Authors:** Emma Pritchard, Karina-Doris Vihta, Koen B. Pouwels, Samuel Lipworth, Russell Hope, Berit Muller-Pebody, T. Phuong Quan, Jack Cregan, Susan Hopkins, David W. Eyre, A. Sarah Walker

## Abstract

**Objective:** Electronic Health Records (EHRs) provide information to explore those at risk of various diseases, though studying entire populations is limited by data availability, potentially introducing biases. We compared different samples, varied by type of hospital contact, to assess the impact on missing data and model results.

**Materials and Methods:** Using *Escherichia coli* bloodstream infections as a case study, we used data from Oxfordshire, UK, containing individuals with hospital contact, including blood tests from primary care providers. We compared two approaches: an “inpatient sample” requiring previous/current inpatient contact, reducing missing risk factors but restricting the denominator, and a broader “healthcare contact sample”, defined by previous healthcare contact (including outpatient appointments, emergency department visits, community blood tests), maximising inclusion.

**Results:** The healthcare contact sample contained more missing data for key demographics and systematically missing data for potential risk factors including diagnosis codes. Missing data was more common in controls than cases (17–21% controls missing vital signs versus 6–7% cases [inpatient sample]) but varied little with lookback duration. Model estimates showed small, insubstantial shifts between the samples. Compared to population estimates, younger males were unrepresented, while those aged≥75y were largely captured.

**Discussion:** Including individuals with any previous healthcare contact resulted in more missing data versus restricting to inpatient-only contact. Despite this, there were only small shifts in model estimates. There were large but expected differences between EHR samples and population-level data.

**Conclusion:** Different sample selection approaches for EHR analyses could impact power and associations. Ideally, multiple approaches should be compared.

## INTRODUCTION

Electronic Health Records (EHRs) offer extensive information to enable identification of high-risk groups for specific diseases (denoted “at-risk” populations). While not their primary purpose, EHRs have been increasingly used for research. The real-world nature of EHR data can reduce costs as data is already collected for routine purposes and allows large sample sizes, useful for rare diseases.[1] EHR use has increased massively over the last 30 years, with 90% of NHS Trusts (hospital groups) in England using EHRs in 2023.[2] Increased linkage between data sources, for example, microbiology samples and hospital admission data nationally in the UK,[3] allows more accurate data on laboratory-confirmed infections and access to more potential risk factors.

When identifying at-risk individuals, ideally, we want to search within the entire underlying population or focus on specific complete populations of interest. However, data availability limitations may restrict this, often with trade-offs between including more individuals in a study and completeness of available data. Restrictions on study populations can introduce missing data and bias estimates of true population risks.

To explore these issues further, here as a case study, we considered identifying risk factors for *Escherichia coli* (*E. coli*) bloodstream infections (BSI). Identifying populations at higher risk of infections can help target interventions to reduce infection rates. As the leading Gram-negative BSI, *E. coli* represents an ideal initial target for reducing infections using linked data to identify associations between risk factors and cases.[4] Reporting of *E. coli* BSIs has been mandatory in the UK since 2011[5] but includes limited risk factors with varying completeness,[6] highlighting the need to use more detailed EHRs.

To investigate risk factors using EHRs, cases and controls (not available in national mandatory surveillance) must be established to compare risk factor differences between cases and a population without the disease of interest (controls). Collider bias can be problematic in studies using EHRs due to the probability of being selected into the study (e.g. healthcare attendance) often being influenced by the exposure and the outcome, inducing incorrect associations (i.e. estimated associations do not have a causal interpretation).[7] This was well documented in hospital-based studies for COVID-19 where, for example, frailty and COVID-19 infection predict hospitalisation.[7] Further, missing data for risk factors can influence results, particularly where multiple imputation is impractical due to study size. Many risk factors are calculable in EHRs from diagnosis codes (ICD-10, 10^th^ revision of the International Classification of Diseases), predominately recorded in inpatient admissions. An absent ICD-10 code may reflect absence of an inpatient admission rather than lack of disease. Missing data and ascertainment bias may therefore be affected by control population selection. Comparing characteristics from multiple samples extracted from the same data source could help assess the impact of sample selection choice on missing data and model results using EHRs. Selection bias into the EHR could be assessed if risk factor prevalence was available in the overall population.

Using data from Oxfordshire, UK, we compared the impact of selecting two different samples, varied by the type of hospital contact, on missing data for potential risk factors and model results. We investigated the impact of lookback duration used to calculate potential risk factors and compared the EHR samples to the general population using Census data and national audits for key potential risk factors.

## METHODS

We used data from the Infections in Oxfordshire Research Database (IORD): a data warehouse including inpatient admissions, outpatient appointments, emergency department (ED) visits, vital signs taken during hospital attendance, and microbiology and biochemistry/haematology results from hospital and primary care samples. Individuals with only primary care visits without blood or microbiology tests, or no primary/secondary care contact, are not represented. IORD includes four large teaching hospitals covering a catchment area of ∼755,000 individuals. The hospital group provides all acute services and laboratory/microbiology testing to the region.

We included all admissions from 1st April 2018-31st March 2024, divided into three two-year blocks, with a five-year lookback, including data from 1st April 2013. Data was analysed separately for the two-year financial year (FY) periods (April to March, keeping winter months together): FY2018/19-2019/20 (1st April 2018-31st March 2020), FY2020/21–2021/22 (1st April 2020-31st March 2022), and FY2022/23–2023/24 (1st April 2022-31st March 2024) presenting results from the most recent two-year period first.

### Case definition

All patients with *E. coli* cultured from blood samples were considered as cases. 69 cultures from individuals<16 years (y) old (as they often have no comorbidities that are common among adults) were excluded (**Figure S1**). One positive blood culture per person was considered in each calendar period, selecting the first *E. coli* BSI if multiple de-duplicated positive cultures were present.

### Control group definitions

As potential controls, we considered all individuals with an inpatient admission, outpatient appointment, ED attendance, or microbiology or blood test recorded in the Laboratory Information Management System (LIMS). The microbiology samples contained both positive and negative results. Records from individuals<16y (196,664 individuals, 1,699,563 records), those missing age (86 individuals, 108 records), and those missing sex (73 individuals, 280 records) were dropped from the analysis (**Figure S2**). 257,362 records (1.3%) from 3,722 (0.4%) individuals with an *E. coli* BSI since 1st April 2013 were removed to reduce control group contamination. If individuals had multiple healthcare visits in a two-year period, only the last contact was included, referred to as the “most recent contact date”.

### Defining analysis samples

Ideally, the control group would include everyone in Oxfordshire; however, IORD includes a subset of individuals with healthcare contact (defined above). Individuals without inpatient episodes may be missing data for many characteristics, risking incorrect classification (ascertainment bias). (Inpatient admissions (spells), are continuous hospital stays from admission to discharge. Each spell contains contiguous consultant (responsible senior doctor) episodes. A new episode begins when the patient transfers to a different consultant or specialty[8]). Two analysis samples were therefore defined based on previous and current healthcare contact in IORD.

The “inpatient sample” included individuals with previous inpatient contact, minimising missing data and ascertainment bias for risk factors. Individuals with *E. coli* BSIs were included if they had a prior inpatient episode ending>72 hours (h) before sample collection date/time defining their *E. coli* BSI (most recent contact date) and in the last 5y (based on episode end date/time). We applied the 72h exclusion to avoid reverse causality. Since many *E. coli* BSIs lead to admission, factors recorded during these episodes may be consequences rather than causes of infection. Controls were included if they had an inpatient episode within the last 5y, ending on or before the “most recent contact” date. The 72h exclusion was not applied as reverse causality was not a concern; however, collider bias was.

The “healthcare contact sample” included individuals with prior healthcare contact with the hospital group, maximising retention for analysis. Individuals with *E. coli* BSIs were included if they had a previous inpatient episode, outpatient appointment, ED visit, blood test, or microbiology/laboratory sample>72h before the collection date/time of BSI and within the last 5y. Controls were included if they had a current or previous inpatient episode (defined above), or a previous outpatient appointment, ED visit, blood test, or microbiology sample strictly before the “most recent contact” date within the last 5y. We excluded current contact from the latter to reduce collider bias by lowering selection based on current health status (demonstrated in **Figure S3**).

### Exposures

We identified six “core variables” selected as key confounders for risk of *E. coli* BSIs: age (calculated at last contact using birth month and year); sex (binary, male/female); ethnicity (binary, white/non-white due to small numbers in the latter); deprivation percentile at recorded residence (Index Multiple Deprivation (IMD), continuous); rural/urban classification at recorded residence (included as a three-level factor); and catchment percentage (percentage of individuals in local area visiting an Oxfordshire hospital as defined by the Office of Health Disparities; 0 = none, 100 = all[9], continuous). See **Supplementary Methods** for risk factor definitions.

### Statistical analyses

We summarised continuous variables as medians with interquartile ranges and categorical variables as proportions. Standardised differences compared core variables, and the amount of missing data, between inpatient and healthcare contact samples for cases and controls.[10]

Presence of *E. coli* BSI (case) versus absence (control) was the binary outcome in all models. We used Poisson models with cluster robust standard errors, as rates were low, making it equivalent to logistic regression but providing a clearer presentation of risk.[11] The “core variables” were included as explanatory variables in all models. Our aim was not to develop a comprehensive risk factor analysis for *E. coli* BSI but to show how associations with “core variables” might change based on sampling strategy. Other risk factors were not included, as these were often unknown for individuals in the healthcare contact sample. Non-linearity was considered for all continuous core variables (**Supplementary Methods**). Incidence-rate ratios with 95% confidence intervals were compared between samples to assess the impact on model estimates.

### Sensitivity Analyses

As one aim when defining the control group was to get close to the Oxfordshire population, we compared the number of individuals captured in IORD to Census data. We compared age and sex distributions for those in the Census with a catchment percentage to Oxfordshire hospitals>80% (due to bimodal distribution, **Figure S4**) with both IORD samples.

People in the true risk set (the entire Oxfordshire population) without comorbidities are likely underrepresented in IORD cohorts, potentially underestimating risk factors if prevalence is lower in the general population. One way to adjust for this is by inflating the dataset using population-level estimates for risk factors, if available. To demonstrate this, we assessed the impact of using IORD versus population-inflated control groups on diabetes risk estimates, a characteristic with available national-level data (**Supplementary Methods**).

Choosing an appropriate lookback period was important for balancing data completeness and relevance. Our main analysis used a five-year lookback (e.g. including individuals who had inpatient contact in the last 5y for the “inpatient sample”) to capture risk factors. However, this may increase missing data due to infrequent healthcare contact or include people living on the border with Oxfordshire attending multiple hospital groups. Therefore, we created additional samples with one to four years of lookback and compared missing data and key demographic model estimates across samples.

## RESULTS

### Data summary

In FY2022/23-2023/24, there were 953 individuals aged≥16y with *E. coli* BSIs confirmed through microbiological isolation. 909 (95%) individuals were in the “healthcare contact sample”, having inpatient, outpatient, ED, microbiology, or blood test contact>72h before BSI collection and within the last 5y (**Figure 1**). 757 (79%) individuals were in the “inpatient sample”, having an inpatient episode ending>72h ago and within the last 5y.

**Figure 1:**
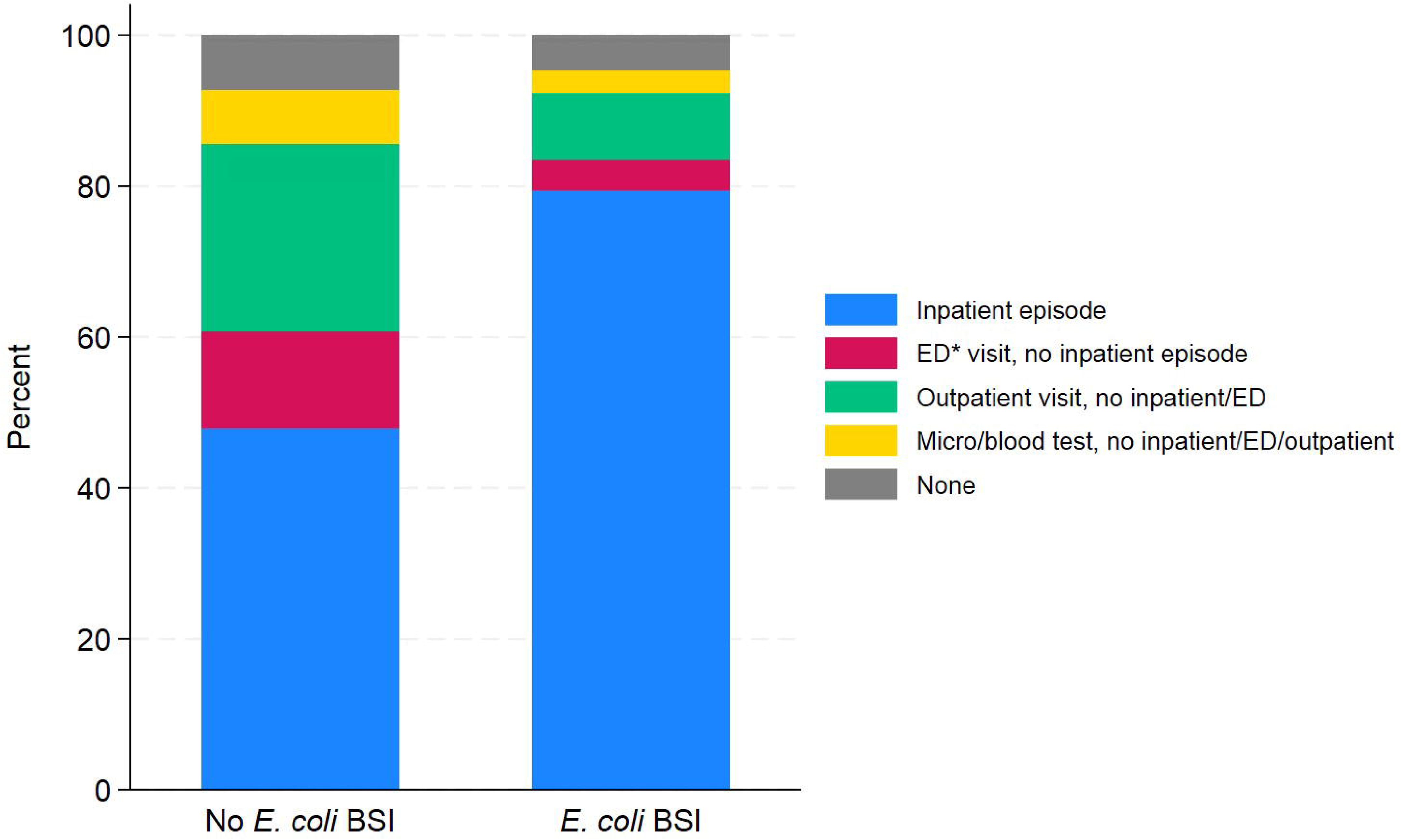
Health contact history in the previous five financial years split by the presence of *E. coli* BSI. *ED = Emergency Department

In FY2022/23-2023/24, there were 577,407 potential controls (76% of IORD catchment population): individuals with inpatient, outpatient, ED, microbiology, or blood test contact within FY2022/23-2023/24. 535,411 (93%) were in the “healthcare contact sample” and 276,758 (48%) in the “inpatient sample” (**Figure 1**). 41,996 (7%) individuals had no current inpatient contact or other contact in the last 5y and were removed from both control groups.

Core characteristics differed between cases and controls, but cases and controls remained similar across the healthcare contact and inpatient samples (standardised differences<0.15, **Table 1**). Cases were generally older than controls, with more women in controls (56%) than cases (48-49%), likely due to maternity-related admissions. Individuals of non-white ethnicities were low in both samples in cases and controls (5%-8% of populations). Missing ethnicity was more common in controls (31% in healthcare contact sample; 165,876/535,411) than cases (16%; 146/909), and higher in the healthcare contact sample than the inpatient sample for both cases (16% vs 13%) and controls (31% vs 23%). The catchment percentage distribution was right-skewed, with most individuals’ home addresses being in high catchment percentage areas (>90%).

**Table 1:** Comparison of the core variables between the healthcare contact sample and the inpatient sample for cases (*E. coli* BSIs) and controls in FY2022/23-2023/24.

|  | Cases, n (%) or median (IQR) |  |  | Controls, n (%) or median (IQR) |  |  |
| --- | --- | --- | --- | --- | --- | --- |
|  | Healthcare contact sample (N=909) | Inpatient sample (N=757) | Standardised difference | Healthcare contact sample (N=535,411) | Inpatient sample (N=276,758) | Standardised difference |
|  | Healthcare contact sample (N=909) | Inpatient sample (N=757) | Standardised difference | Healthcare contact sample (N=535,411) | Inpatient sample (N=276,758) | Standardised difference |
| Age (years) | 77 (65, 86) | 78 (66, 86) | 0.10 | 55 (36, 69) | 57 (38, 73) | 0.14 |
| Missing | 0 (0) | 0 (0) |  | 0 (0) | 0 (0) |  |
| Sex |  |  |  |  |  |  |
| Male | 465 (51) | 396 (52) | 0.02 | 237,448 (44) | 120,740 (44) | 0.01 |
| Female | 444 (49) | 361 (48) |  | 297,963 (56) | 156,018 (56) |  |
| Missing | 0 (0) | 0 (0) |  | 0 (0) | 0 (0) |  |
| Ethnicity |  |  |  |  |  |  |
| White | 707 (78) | 612 (81) | 0.01 | 330,784 (62) | 190,546 (69) | 0.01 |
| Non-white | 56 (6) | 46 (6) |  | 38,751 (7) | 21,489 (8) |  |
| Missing | 146 (16) | 99 (13) |  | 165,876 (31) | 64,723 (23) |  |
| Deprivation percentile | 74 (54, 89) | 74 (54, 89) | 0.01 | 74 (54, 89) | 74 (54, 89) | -0.01 |
| Missing | 8 (1) | 4 (1) |  | 6310 (1) | 2567 (1) |  |
| Rural/urban classification |  |  |  |  |  |  |
| Urban area | 601 (66) | 498 (66) | 0.02 | 349,611 (65) | 181,153 (65) | 0.01 |
| Rural town | 152 (17) | 132 (17) |  | 85,077 (16) | 44,813 (16) |  |
| Rural village | 148 (16) | 123 (16) |  | 94,402 (18) | 48,219 (17) |  |
| Missing | 8 (1) | 4 (1) |  | 6,321 (1) | 2,573 (1) |  |
| Catchment percentage | 95 (88, 96) | 95 (89, 96) | 0.11 | 93 (40, 96) | 93 (62, 96) | 0.06 |
| Missing | 8 (1) | 4 (1) |  | 6,310 (1) | 2,567 (1) |  |

*Note: For deprivation percentile, 1=most deprived, 100=least deprived. Catchment percentile is the percentage of individuals in the local area visiting an Oxfordshire hospital as defined by the Office of Health Disparities; 0 = none, 100 = all.[9]*

There was less missing data for potential risk factors in cases than controls, as more cases had previous inpatient contact. Missing data was higher in the healthcare contact sample versus the inpatient sample, particularly for blood tests and vital signs (**Figure 2**). The amount of missing data varied between blood tests as some, like HbA1c, may not be routinely done (19% cases and 40% controls missing HbA1c in the healthcare contact sample). More individuals in the healthcare contact sample had missing results for routine blood tests, such as haemoglobin, neutrophils, or lymphocytes; 8% of controls in the inpatient sample were missing haemoglobin versus 19% in the healthcare contact sample. Prostate-specific antigen (PSA) was missing in large numbers of cases (45-50% of men) and controls (70-75% of men) in both samples with small differences between samples (standardised difference 0.07 and 0.06 for cases and controls, respectively). There were large differences in the percentage of individuals missing urea tests; 20% in inpatient sample controls versus 45% in healthcare contact sample controls (standardised difference=0.54). Few cases were missing weight and body mass index (BMI) measurements. Controls had more missing data, particularly in the healthcare contact versus the inpatient sample (38% versus 13% missing weight; 62% versus 40% missing BMI).

**Figure 2:**
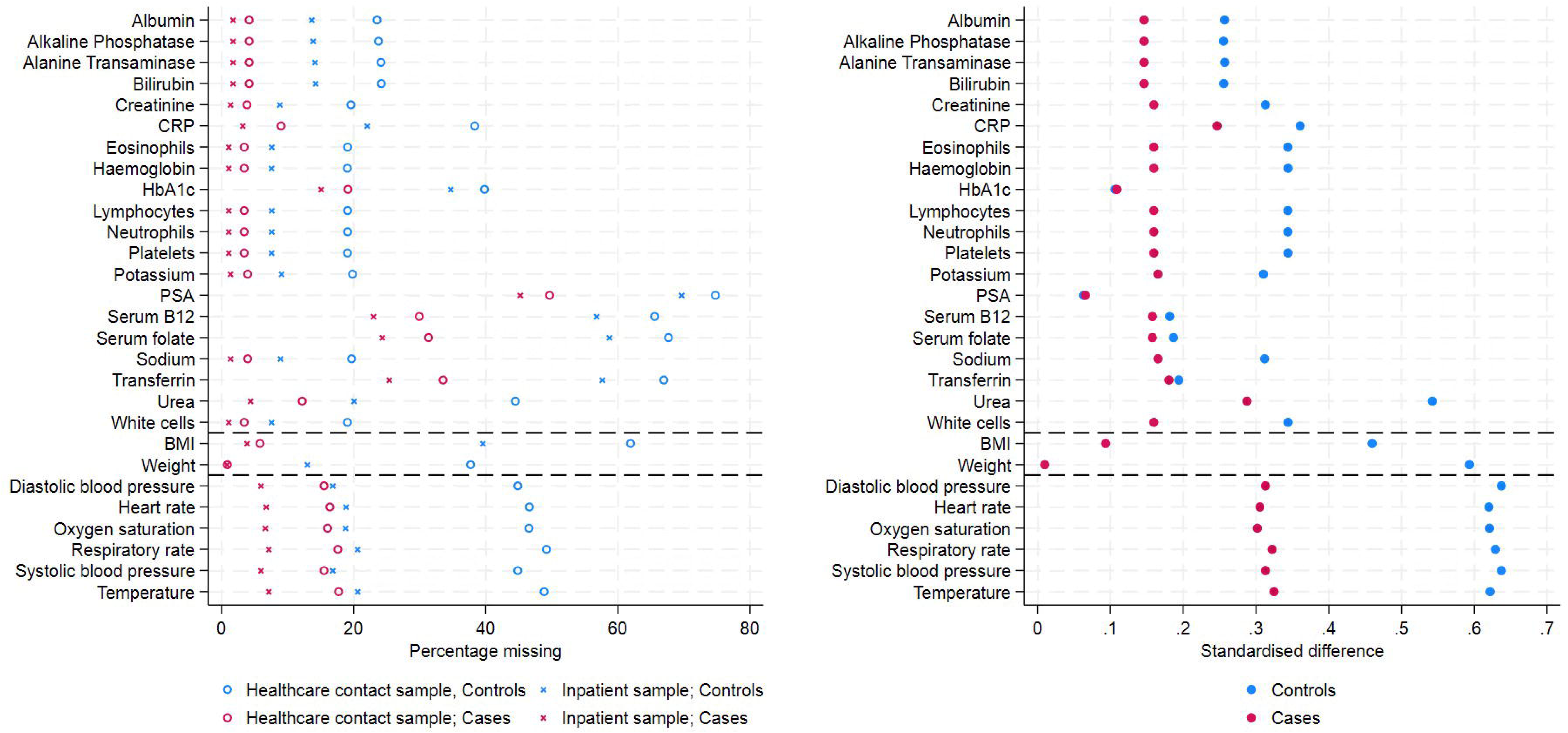
Percentage of missing data for potential risk factors (left), split by healthcare contact versus inpatient samples and the presence of *E. coli* bloodstream infection, and standardised difference (right) between the proportion missing in the inpatient and healthcare contact samples separately for cases and controls. Note: Horizontal black dashed lines separate blood tests (top section), personal characteristics (middle section) and vital signs (bottom sections). CRP=C-reactive protein; PSA=prostate-specific antigen. The denominator for PSA tests is men only as the test is only done in men.

Some potential risk factors were systematically missing, only recorded in inpatient admissions or ED attendances, e.g. vital signs. 6-7% of cases in the inpatient sample were missing vital sign measurements, compared with 16-18% in the healthcare contact sample (standardised differences 0.30-0.32) (**Figure 2**). More controls were missing vital signs measurements: 17-21% in the inpatient sample and 45-49% in the healthcare contact sample (standardised difference between 0.62-0.64). While diagnosis and procedure codes are recorded in some outpatient appointments, this was not routine for all attendances; 7% (971,182/13,495,823) of outpatient appointments between 1st April 2013-31st March 2024 (the study period plus 5y lookback) had at least one diagnosis code, versus 98% (2,504,887/2,544,974) of inpatient episodes. For procedure codes, 13% (1,818,563/13,495,823) of outpatient appointments versus 65% (1,658,093/2,544,974) of inpatient episodes had at least one procedure code.

### Comparison of model results

Despite variation in size of control populations, there were few differences between estimates from Poisson models including the core variables using the different samples (**Figure 3**). In both samples, older age was associated with higher risk of *E. coli* BSIs. Non-white ethnicity was associated with higher risk of *E. coli* BSIs, however, this was not significant at p<0.05. Being female was similarly associated with decreased risk of *E. coli* BSIs compared with males in both samples; inpatient sample IRR 0.80 (95% CI 0.69-0.93), healthcare contact sample IRR 0.83 (95% CI: 0.72-0.95). Lower deprivation was associated with lower *E. coli* BSI risk and higher catchment percentage with higher *E. coli* BSI risk, likely due to improved ascertainment, however, this attenuated slightly in the healthcare contact sample. Living in rural settings was associated with lower *E. coli* BSI risk versus living in urban cities/towns.

**Figure 3:**
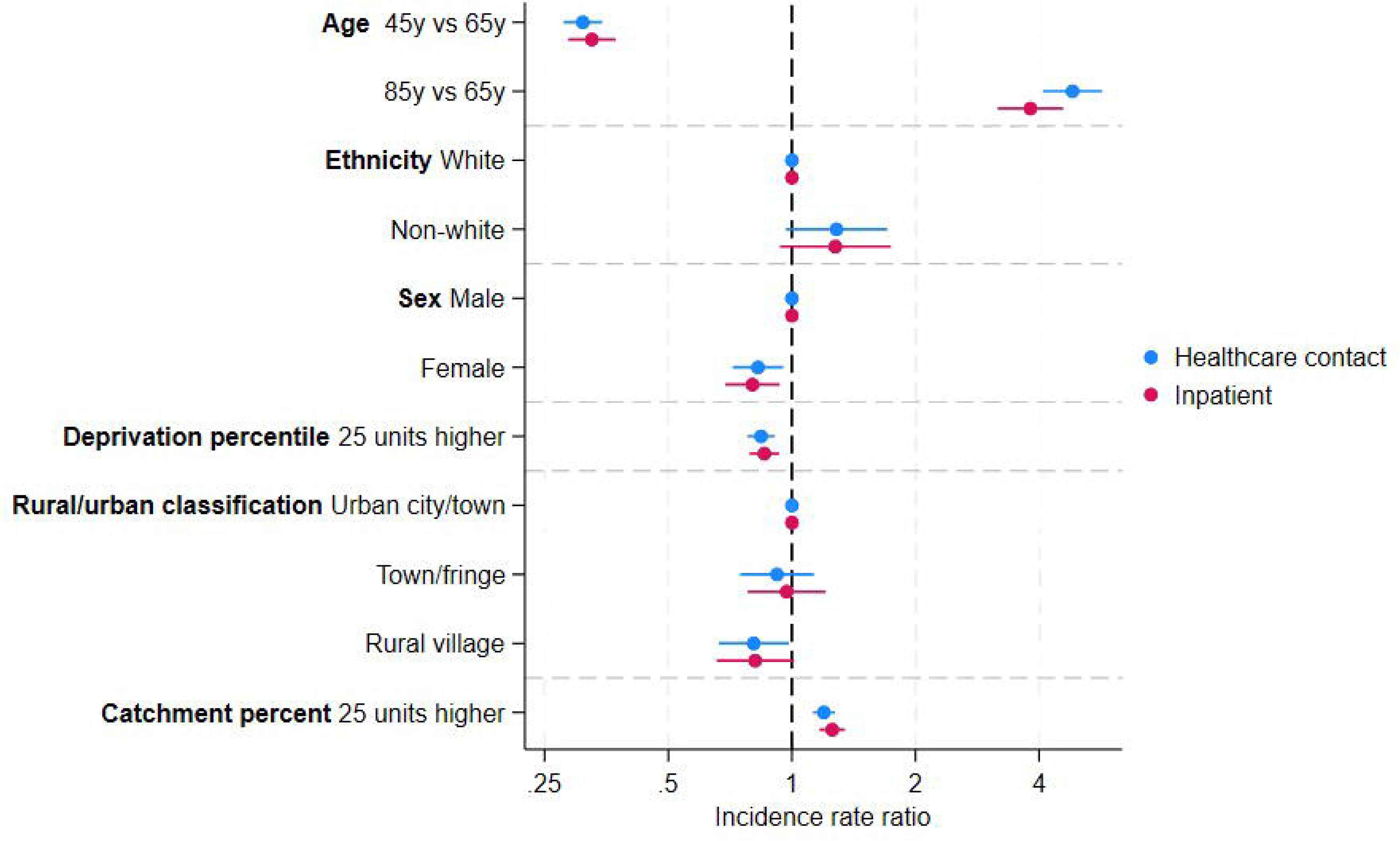
Incidence rate ratios with 95% confidence intervals for core variables from Poisson models run on the healthcare contact sample (blue) and the inpatient sample (red) separately. Note: Age was included as a restricted cubic spline (one internal knot) with the values presented predicted from the fitted spline. For deprivation percentile, 1=most deprived, 100=least deprived. Catchment percentile is the percentage of individuals in the local area visiting an Oxfordshire hospital as defined by the Office of Health Disparities; 0 = none, 100 = all.[9]

### Comparison to population-level data

We observed large but expected differences in age and sex distributions comparing individuals in the IORD samples to the Office for National Statistics census data for Oxfordshire (**Figure 4**). The largest difference was in the youngest age groups (16-24y), particularly for the inpatient sample: 38,215 men were recorded in the census versus 7,575 (20%) in the inpatient sample (36,670 versus 10,772 for women). The number of women aged 25-34 in the healthcare contact sample was similar to the census (43,868 versus 44,488), decreasing to 31,622 when only including those with >80% catchment in IORD. This age group was underrepresented in men (24,816 versus 43,993). Older age groups were captured more completely, with the inpatient sample including a similar number of people for those aged≥75y, with more individuals than the census included in the healthcare contact. Including those with ≤80% catchment contributed to this overrepresentation e.g. 20,201 women aged 75-84y in the census; 19,778 in the healthcare contact with >80% catchment and 8,746 with ≤80% catchment.

**Figure 4:**
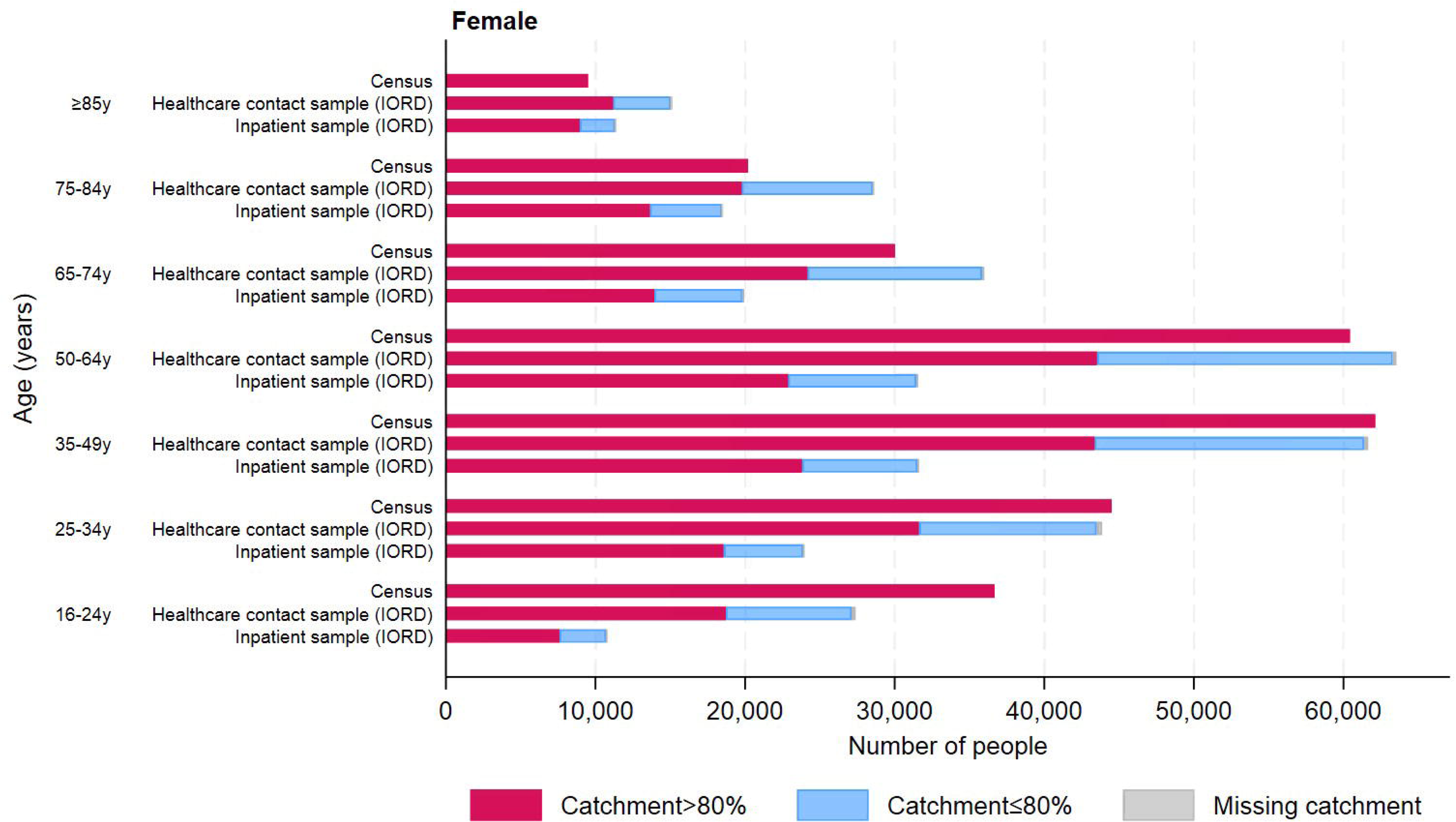
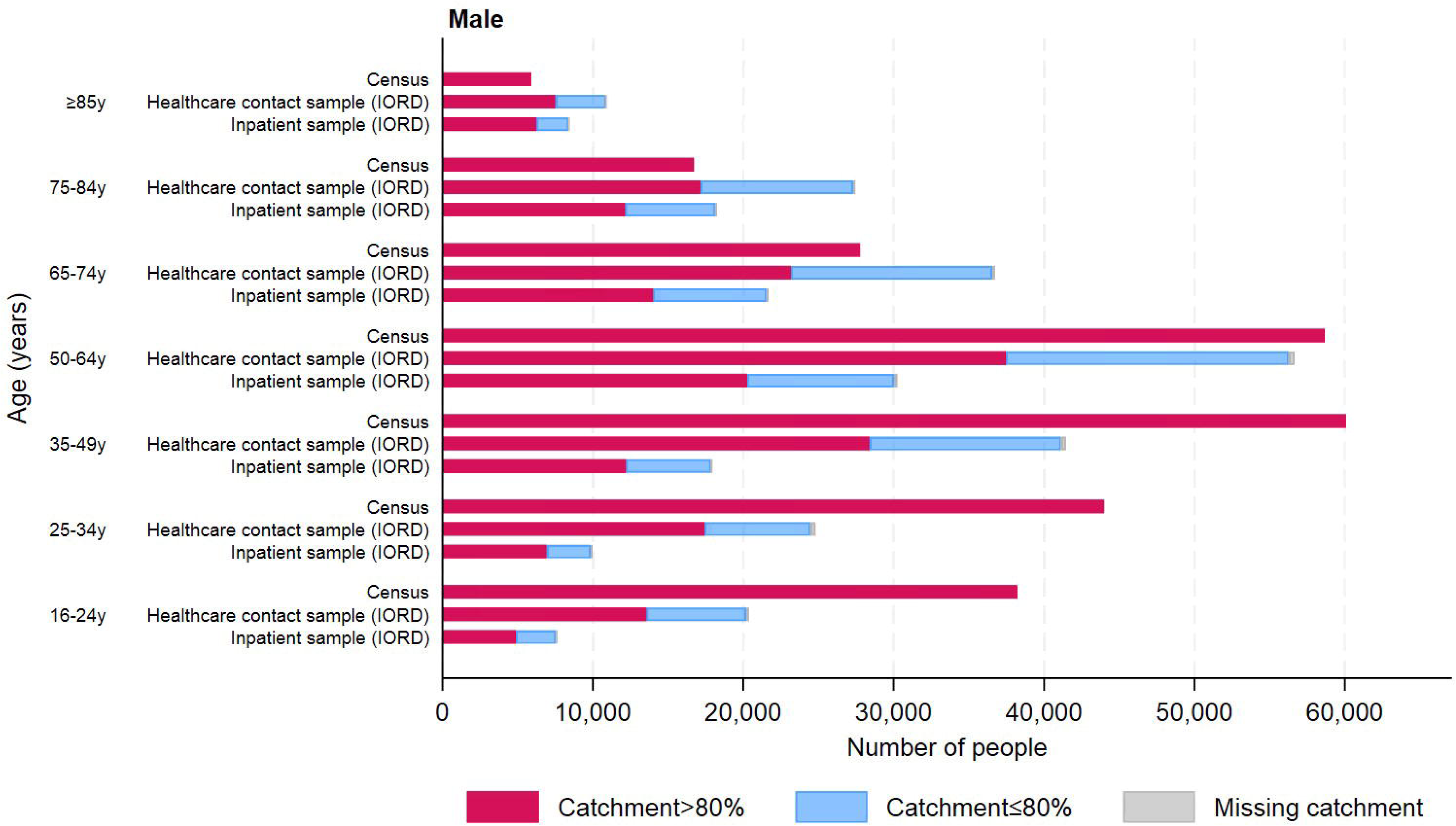
Comparison of age and sex distributions between the healthcare contact sample, inpatient sample from FY2020/21-2021/22, and the general population using 2021 census data from the Office for National Statistics.

For an example risk factor of diabetes, we found that the number of diabetes cases in IORD closely matched population-level data for those aged≥80 (median difference: -1 [IQR: -8, 5] cases per primary care practice). However, there were more cases in population-level data for those aged 65-79y and 40-64y. Similar patterns were observed for those registered in primary care but without diabetes (**Figure S5-S6, Table S1**). Inflating the control group to match population levels increased control group size, while slightly attenuating model effects (**Tables S2-S4; Supplementary Results**).

### Comparison of lookback lengths

Longer lookback length increased the number of cases and controls, especially in the inpatient sample (**Table 2**). Using only 1y lookback decreased the case and control numbers by 20% (n=604) and 46% (n=153,902) versus 5y lookback, respectively. An additional 76 cases (13% percentage increase) and 45,953 controls (30% percentage increase) were added when increasing from 1y-2y lookback. The number of cases and controls added with each increasing year of lookback decreased as lookback length increased; however, there were an additional 23,304 (9%) controls added when increasing from 4y-5y lookback in the inpatient sample. There was little benefit when increasing lookback length beyond 2y in the healthcare contact sample.

**Table 2:** Number of cases and controls in the inpatient and healthcare contact samples with one to five years of lookback length in the FY2022/23-2023/24 samples.

| Lookback length (years) | Inpatient sample |  | Healthcare contact sample |  |
| --- | --- | --- | --- | --- |
|  | *E. coli* cases, n (%) | Controls, n (%) | *E. coli* cases, n (%) | Controls, n (%) |
| 1 | 604 (80) | 153,902 (56) | 877 (96) | 465,327 (87) |
| 2 | 680 (90) | 199,855 (72) | 892 (98) | 506,471 (95) |
| 3 | 711 (94) | 230,892 (83) | 899 (99) | 520,402 (97) |
| 4 | 744 (98) | 253,454 (92) | 905 (100) | 529,096 (99) |
| 5 | 757 (100) | 276,758 (100) | 909 (100) | 535,411 (100) |
| Potential* | 953 | 577,407 | 953 | 577,407 |

\*Potential number of cases with no lookback restrictions. Percentages are given using the five-year lookback populations.

For some blood tests, e.g. HbA1c and albumin, increasing lookback length decreased missing data in cases and controls in the inpatient sample, whilst increased lookback in the healthcare contact sample increased missing data, suggesting individuals being added into the control groups with increasing lookback were more and less likely to have these variables measured, respectively (**Figure 5**). This may be because these blood tests are more common during inpatient admissions, so including individuals with previous admissions increases the likelihood of capturing these measurements. Missing data increased as lookback length increased for vital signs (only recorded in inpatient admissions and ED visits); adding previous outpatient or ED visits may increase missing data as these tests are less likely to have been done in their previous, non-inpatient, contact.

**Figure 5:**
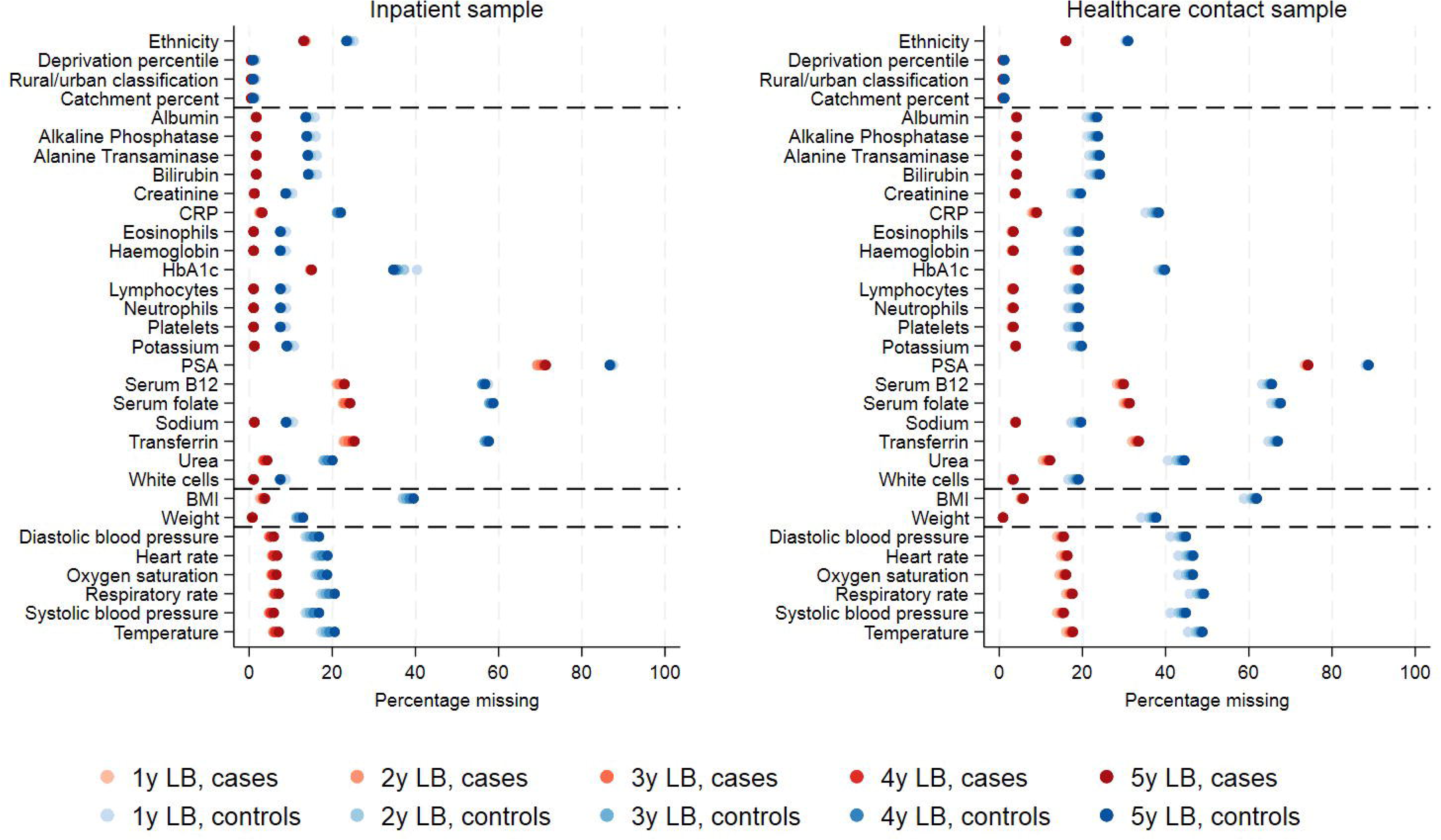
Percentage of missing data for core variables and potential risk factors in *E. coli* BSI cases and controls in FY2022/23-2023/24 with varying lookback. Note: Horizontal black dashed lines separate core variables (top section), blood tests, personal characteristics, and vital signs (bottom sections). CRP=C-reactive protein; PSA=prostate-specific antigen.

Estimates from the core model using different lookback lengths varied slightly for both the inpatient and healthcare contact samples; however, interpretations from the models remained similar (**Figure 6**). The largest difference was attenuation effects for older ages as lookback length decreased in the inpatient sample, perhaps reflecting a higher proportion of older individuals with recent inpatient admissions.

**Figure 6:**
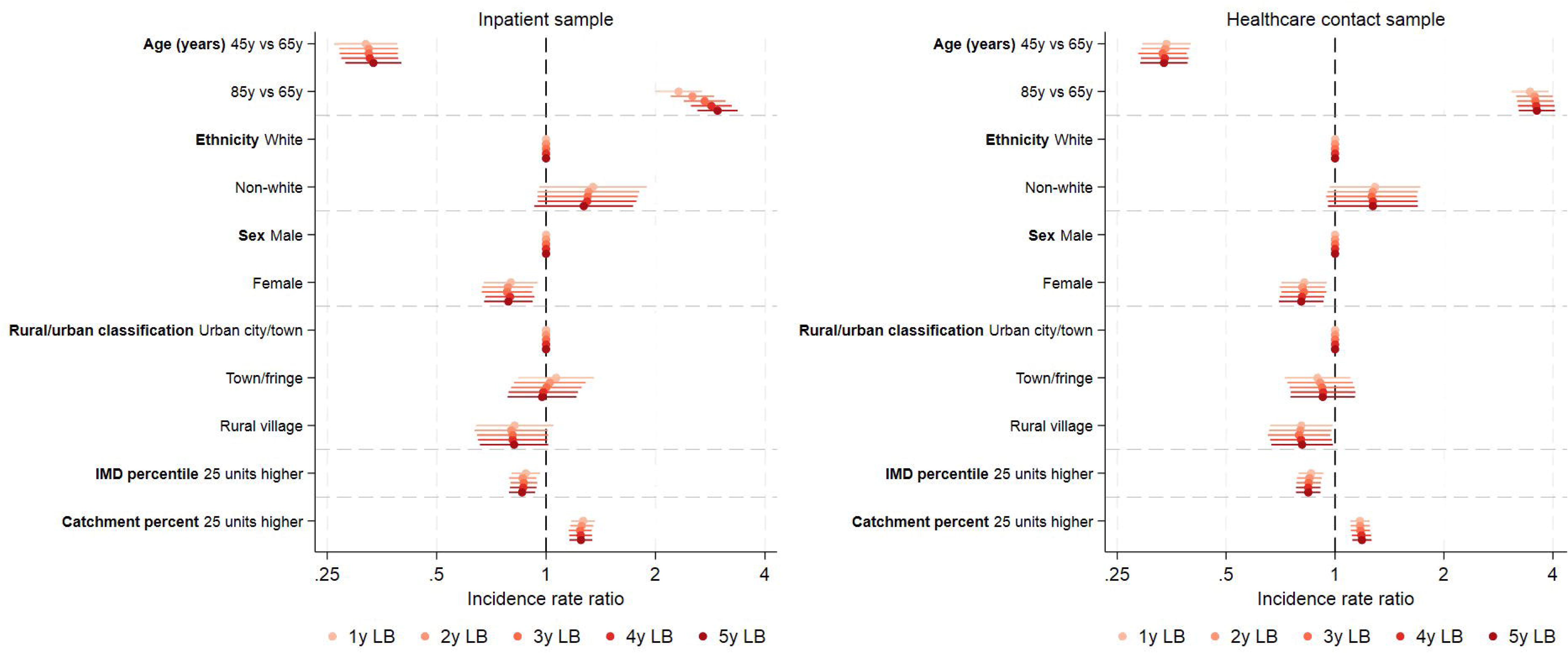
Model estimates comparing different lookback lengths in the inpatient (left) and healthcare contact (right) samples in FY2022/23-2023/24.

Similar results were observed in previous periods, specifically FY2018/19-2019/20 and FY2020/21-2021/22 (**Supplementary Results**, **Table S5**, **Figures S7-S11**).

## DISCUSSION

Using EHRs, we investigated how different sample selections impacted estimated associations between *E. coli* BSIs and key demographics and missingness of potential risk factors. We found that including individuals with any previous healthcare contact (“healthcare contact” sample, including inpatient, outpatient, ED, microbiology, or blood test contact) resulted in more missing data for core characteristics, such as ethnicity, and potential risk factors. For example, only vital signs recorded in inpatient admissions and ED visits were available in IORD. Despite this, there were only small shifts in model estimates for core variables between the inpatient and healthcare contact samples. There were expected differences in demographics comparing IORD to the Census. Missingness of variables varied by lookback length, with missing data both increasing and decreasing with increased lookback for different variables.

Choosing who to include in control groups was a balance between getting close to the target population of Oxfordshire while reducing missing data in risk factors (and ascertainment bias in risk factors based on diagnosis/procedure codes) and avoiding inclusion of individuals unlikely to visit Oxfordshire hospitals for an *E. coli* BSI, reducing ascertainment bias in the outcome. One limitation is using data from one hospital group in England missing attendances elsewhere, especially for patients near Oxfordshire’s border. An estimated 25% of individuals with multiple hospital encounters in England (2017-2018) visited ≥two hospital groups, with regional variations.[12] While percentages per hospital were not reported, 54% of those attending multiple Trusts visited 20 hospital pairs, none involving Oxfordshire hospitals, suggesting limited impact on IORD data. Patients residing outside Oxfordshire may attend Oxford University Hospitals, e.g. when travelling into the area for holidays, or attending specialist care e.g. cardiac specialist services.[13] *E. coli* BSIs from these patients may therefore be missed. We attempted to account for this variation in healthcare attendance by requiring individuals to have contact in the current year and adjusting for catchment percentage.

While proving collider bias has not impacted model results is generally not possible,[7] we attempted to assess its potential impact by comparing the age/sex distribution to the target population of Oxfordshire and inflating the control group to match national diabetes estimates. Having an unrepresentative sample and drawing inferences to larger populations can introduce bias.[14] We compared model estimates between the “inpatient” and “healthcare contact” samples to assess whether selection bias influenced inpatient sample results, showing little difference between model estimates of key demographics. As most individuals (79%) with *E. coli* BSIs had previous inpatient episodes in the last 5ys, fewer cases were removed from analysis, compared with 48% of controls having inpatient contact. This may reduce, but not eliminate, the risk of collider bias. Studying diseases which do not almost exclusively result in hospitalisation, or where previous hospital contact is rarer, may have a larger impact on model results and introduce more selection bias. For example, if studying risk factors for COVID-19 in EHRs, analysis will be restricted to those hospitalised with COVID-19 which is a much smaller subset[7] (estimated 2.1% of people with infection[15]). When considering selection bias, it may be more appropriate to interpret risk factors conditional on the sample used. For example, risk factors found in the “inpatient sample” reflect risk factors within the sub-population of individuals with an inpatient admission in the last 5y. This may still be practically useful for targeting interventions.

Using EHR data risks missing individuals without contact with healthcare facilities. We demonstrated how to inflate datasets based on population-level estimates, using diabetes as an example, however, this approach has limitations. Population-level estimates for most risk factors are unavailable, with diabetes being an exception. Even when such data exists, definition differences can pose challenges. Additionally, other characteristics must be imputed. In our analysis, we assumed added individuals had the same age/sex distribution as observed in the EHR conditional on diabetes, but unobserved individuals may differ due to health-seeking behaviour. While this approach is feasible, it relies on strong assumptions making it challenging to implement.

There was more missing data in the healthcare contact versus the inpatient sample, with data often being systematically missing. Ethnicity was missing between 20-27% of individuals in the control groups, higher than 13-17% observed in national-level data in 2019/20.[16] Another UK-based study found younger, male individuals with fewer comorbidities had higher missing ethnicity, likely due to less contact with the healthcare system, decreasing opportunities to record data.[17] Other potential reasons include reduced opportunities for staff to ask patients about ethnicity or lack of knowledge of why this data collection is important.[18] While ICD-10 and procedure codes are used for billing purposes, outpatient appointments are billed via clinical codes, making ICD-10 and procedure codes optional and sporadically recorded.[19] Missing data varied for blood test results, with some tests carried out routinely, while others conducted upon suspicion of illness, e.g. HbA1c for suspected or confirmed diabetes. Certain blood tests may also be age-dependent; e.g. with prostate cancer being rare in men aged<50y, PSA testing is often done on men aged≥50y.[20]

The main limitation was conducting this study using a single population from Oxfordshire. While this was a large sample accounting for ∼1% of the UK population, generalisability may be limited; future research could explore national linked data. Further, there was no access to primary care records meaning, for example, only vital signs taken in inpatient admissions and ED visits could be included. Access to primary care records may have allowed a larger proportion of the Oxfordshire population to be studied and more risk factors defined, however, linked primary and secondary care, and microbiology data, are not yet available at scale. Lastly, we only considered one outcome; however, with the large number of controls, exposure distributions are likely similar to other populations. Assessing the impact of the different samples would be important if studying different diseases; however, the framework established in this paper may offer a pragmatic way to do so.

## CONCLUSION

Overall, consideration has to be taken when defining case and control groups and sample selection in studies using EHRs to avoid introducing bias. Future work focusing on the impact of the sample selection on other diseases, and exploring impact in national data, may be helpful going forward.

## Acknowledgements

This work uses data provided by patients and collected by the UK’s National Health Service as part of their care and support. We thank all the people of Oxfordshire who contribute to the Infections in Oxfordshire Research Database. Research Database Team: L Butcher, H Boseley, C Crichton, DW Crook, D Eyre, O Freeman, J Gearing (community), R Harrington, K Jeffery, M Landray, A Pal, TEA Peto, TP Quan, J Robinson (community), J Sellors, B Shine, AS Walker, D Waller. Patient and Public Panel: G Blower, C Mancey, P McLoughlin, B Nichols.

## Author contributions

The analysis was designed and planned by ASW, DWE, SH, and EP. EP analysed the data and created the visualisations. EP, ASW, and DWE drafted the manuscript and all authors contributed to the interpretation of the data and results and revised the manuscript. All authors approved the final version of the manuscript.

## Data availability

The datasets analysed during the current study are not publicly available as they contain personal data but are available from the Infections in Oxfordshire Research Database (https://oxfordbrc.nihr.ac.uk/research-themes-overview/antimicrobial-resistance-and-modernising-microbiology/infections-in-oxfordshire-research-database-iord/), subject to an application and research proposal meeting the ethical and governance requirements of the Database. For further details on how to apply for access to the data and for a research proposal template please.

