## Supplementary Material for "The effect of population selection criteria on model estimates and data missingness in electronic health record studies"

### Supplementary Methods

**Defining exposures**

Potential risk factors were defined from all available data sources to assess missingness across the two samples; specifically inpatient admissions and outpatient appointments including ICD-10 codes and procedure codes (OPCS Classification of Interventions and Procedures version 4), ED attendances (which do not have these codes but a different coding system for recording reasons for attendance which is not aligned with ICD-10 and hence not used), blood test results, microbiology results, and vital signs. Risk factors were selected based on pre-defined groupings (e.g. Summary Hospital-level Mortality Indicator groupings of ICD-10 codes[1]), previously published research (e.g.[2, 3]), clinical advice and knowledge, and the availability of data. A full list of risk factors and definitions is available at: https://github.com/EmmaPritchard/EHR-Risk-Factor-Definitions.

Risk factor definitions did not include variables recorded in the 72 hours directly before the blood culture collection date/time for *E. coli* cases to avoid issues of reverse causality. Risk factors calculated for the control group were not subject to this exclusion and hence all variables contributing to each risk factor recorded in the previous five years up to and including the current contact were considered in calculations. Blood tests taken outside of hospital were prioritised because they were more likely to reflect a "steady state" than acute illness, followed by measurements at outpatient appointments, measurements closest to discharge from an inpatient admission, and lastly, measurements taken at ED visits. The same priority was used for vital signs, but no measurements were available if taken outside of the hospital setting.

**Defining non-linearity in models**

Restricted cubic splines with between one to five internal knots were considered for all continuous core variables, selecting the parameterisation based on the Bayesian information criterion. A minimum of one internal knot was included for age due to expected variation. Internal knots were placed at even percentiles throughout the range of values for each variable (after truncation at the 5th and 95th percentiles) with boundary knots included at the 10th and 90th percentiles.

**Inflating the control group to match population-level estimates**

Additional diabetes cases were obtained from the National Diabetes Audit (NDA),[4] which reports type 2 diabetes prevalence by GP practice across four age groups (<40y [including children], 40–64y, 65–79y, and 80y+). Since individuals aged <16 were excluded from the main analysis, only the three older age groups were considered.

To add additional controls without diabetes, we used the total number of people registered at each GP practice (from NHS Digital)[5] and subtracted the diabetes cases recorded in the NDA. The number of *E. coli* BSI cases with and without diabetes remained unchanged, based on the assumption that all *E. coli* BSIs in Oxfordshire were captured in the dataset and that diabetes was recorded for all individuals with *E. coli* BSIs with diabetes.

This analysis was conducted on a subset of individuals registered at a GP practice with >80% catchment to Oxford University Hospitals Trust (OUH), as *E. coli* cases from outside this range are less likely to be captured, which would be incompatible with the assumption that all individuals with *E. coli* BSIs with diabetes have been captured.

To impute missing values for sex and ethnicity in the added rows, we assumed they followed the same distribution as in the observed IORD data, conditional on diabetes and age. Specifically, we fitted separate logistic regression models for sex and ethnicity for each age group, using diabetes status as the sole predictor. Predicted probabilities were generated from the models, and random draws from a uniform distribution were used to assign binary outcomes. While rural/urban classification was included in the core model, it was excluded here as there was little evidence of an effect in the main analysis. Catchment percentage and age were also not imputed as the data was restricted to individuals with catchment >80% and conducted separately by age groups.

Model estimates from Poisson models estimating the effect of diabetes on the presence of *E. coli* BSI were compared between the IORD dataset and the population-inflated datasets, with models fit separately for each age group and adjusted for sex and ethnicity.

### Supplementary Results

**Inflating the control group to match population-level estimates**

There was a total of 62 GP practices with a catchment percentage of >80% catchment for Oxford University Hospitals (OUH) Trust, with 738,417 individuals of all ages registered to these GPs (from NHS digital) and 28,860 individuals with type 2 diabetes at these GPs (overall 3.9% prevalence, NDA). In IORD, these GP practices covered 17,219/24,173 (71%) of all type 2 diabetes cases in the FY2020/21-2021/22 sample.

The number of individuals with type 2 diabetes recorded in the IORD differed from that in the NDA, with variations observed across age groups (**Figure S5**, **Supplementary Tables**

Table S1). For individuals aged 80+, the number recorded in the IORD closely matched the NDA, with a median difference of -1 person (IQR: -8, 5) across the 62 GP practices with >80% catchment for OUH. In contrast, there was a median of 60 more individuals (IQR: 34, 89) with diabetes in the 65–79-year age group in the NDA versus IORD, increasing to 100 more individuals (IQR: 54, 156) in the 40–64-year age group.

A similar trend was observed for individuals without diabetes but registered at the GP, with national data showing a median of 1,425 additional individuals (IQR: 827, 2,043) compared to IORD for the 40–64-year age group (**Figure S6**, **Supplementary Tables**

Table S1). However, for the 80+ age group, IORD data included a median of 35 more individuals per GP practice than the corresponding national-level data.

Inflating the datasets to match national-level data, an additional 95,168 individuals (78% relative increase) were included as never having diabetes in the controls, and 6,899 (135% relative increase) as having diabetes in the controls for those aged 40-64 (**Table S2**). Relatively, fewer individuals were added for the 65-79y age group; 12,640 (20% increase) more individuals without diabetes, and 4,067 (61% increase) with diabetes (**Table S3Error! Reference source not found.**).

In both age groups, there was a slight attenuation of the associated effect of diabetes on the risk of *E. coli* BSIs in Poisson models adjusted for sex and ethnicity (**Table S4**).

**Results from different periods**

While the most recent period was of most interest when trying to identify risk factors, we wanted to assess whether results were consistent across other periods. We therefore evaluated the impact of the different sample selections on the two previous non-overlapping periods: FY2020/21-2021/22 (1st April 2020-31st March 2022) and FY2018/19-2019/20 (1st April 2018-31st March 2020).

Similarly to FY2022/23-2023/24, a small proportion of controls (7% FY2020/21-2021/22 and 8% FY2018/19-2019/20) and cases (4% FY2020/21-2021/22 and 4% FY2018/19-2019/20) had no current inpatient contact, and no other contact in the last 5ys, excluding them from both the inpatient and healthcare contact sample (**Figure S7**). Around 50% of controls and 80% of cases were in the inpatient sample in all three periods. There was little difference in core variables between the healthcare contact and inpatient samples within cases and controls across the different periods, while characteristics varied consistently between cases and controls (**Table S5**).

There was more missing data for vital signs in FY2018/19-2019/20 than in other periods, with 55-58% controls and 24-25% cases missing measurements in the healthcare contact sample, compared with 49-52% controls and 18-20% of cases in FY2020/21-2021/22 due to electronic recording of vital signs only consistently being recorded in IORD from January 2016 onwards (**Figure S8**). Again, there was more missing data for all potential risk factors in the healthcare contact sample compared with the inpatient sample, and more missing data in controls than cases.

There were few differences between estimates from Poisson models including the core variables in both periods, with a slight attenuation of age in the inpatient sample compared with the healthcare contact sample (**Figure S9**). The effect of catchment percentage was smaller in FY2020/21-2021/22 (1.15 [95% CI 1.08, 1.22]) and FY2022/23-2023/24 (1.20 [95% CI 1.13, 1.27]) compared with FY2018/19-2019/20 (1.33 [95% CI 1.24, 1.43]).

Again, longer lookback lengths both increased and decreased the amount of missing data of core characteristics and potential risk factors (**Figure S10**). The largest difference in proportion missing was increasing from 1y to 5y lookback in the control group of the inpatient sample in FY2018/19-2019/20 e.g. 25% missing temperature to 34% missing temperature, respectively. In comparison, 19% were missing temperature using 1y lookback for controls in FY2020/21-2021/22 and 24% using a 5y lookback. All other missingness patterns were similar to the FY2022/23-2023/24 period, with most blood tests having less missing data with increased lookback in the inpatient sample but more missing data with increased lookback in the healthcare contact sample.

Estimates from the core model using different lookback lengths varied slightly in the inpatient and healthcare contact samples, however, interpretations of model results remained similar (**Figure S11**).

### Supplementary Tables

**Table S1: Median (IQR) difference in the number of people with and without diabetes registered at primary care practices versus available in IORD by age group.**

| Age group (years) | Difference in the number of people with type 2 diabetes, median (IQR) | The difference in the number of people without type 2 diabetes, median (IQR) |
|---|---|---|
| 40-64 | 100 (54, 156) | 1,425 (827, 2,043) |
| 65-79 | 60 (34, 89) | 175 (97, 262) |
| 80+ | -1 (-8, 5) | -35 (-56, -20) |

**Table S2: Number of individuals aged 40-64y in IORD, split by diabetes status and presence of *E. coli* BSI. Inflated controls were estimated from the National Diabetes Audit and primary care registration data.**

| Type 2 diabetes | Original controls, n (%) [N=127,579] | Inflated controls [N=229,646] | *E. coli* BSI, n (%) [N=110] |
|---|---|---|---|
| **>5y ago or never in EHR** | 122,489 (96) | 217,657 (95) [+95,168, 78% increase] | 88 (80) |
| **≤5y ago** | 5,090 (4) | 11,979 (5) [+6,899, 135% increase] | 22 (20) |

*Note: Only individuals registered at primary care practices with >80% catchment were included.*

**Table S3: Number of individuals aged 65-79y in IORD, split by diabetes status and presence of *E. coli* BSI. Inflated controls were estimated from the National Diabetes Audit and primary care registration data.**

| Type 2 diabetes | Original controls, n (%) [N=70,956] | Inflated controls [N=91,730] | *E. coli* BSI, n (%) [N=110] |
|---|---|---|---|
| **>5y ago or never in EHR** | 64,280 (91) | 76,920 (88) [+12,640, 20% increase] | 159 (74) |
| **≤5y ago** | 6,676 (9) | 10,743 (12) [+4,067, 61% increase] | 57 (26) |

*Note: Only individuals registered at primary care practices with >80% catchment were included.*

**Table S4: Incidence rate ratios for the association between diabetes and *E. coli* BSIs from Poisson models including diabetes and adjusted for sex and ethnicity.**

| Dataset | Age group | IRR (95% CI) | P-value |
|---|---|---|---|
| **Original dataset** | 40-64 | 6.4 (3.9, 10.5) | <0.001 |
| **Inflated dataset** | 40-64 | 5.0 (3.0, 8.2) | <0.001 |
| **Original dataset** | 65-79 | 3.2 (2.3, 4.4) | <0.001 |
| **Inflated dataset** | 65-79 | 2.4 (1.7, 3.3) | <0.001 |

**Table S5: Comparison of the core variables between the healthcare contact sample and the inpatient sample for cases (*E. coli* BSIs) and controls in FY2020/21-2021/22 and FY2018/19-2019/20.**

| | FY2020/21-2021/22 | | | | | | FY2018/19-2019/20 | | | | | |
|---|---|---|---|---|---|---|---|---|---|---|---|---|
| | Cases, n (%) or median (IQR) | | | Controls, n (%) or median (IQR) | | | Cases, n (%) or median (IQR) | | | Controls, n (%) or median (IQR) | | |
| | Healthcare contact sample (N=818) | Inpatient sample (N=677) | SD* | Healthcare contact sample (N=493,676) | Inpatient sample (N=260,977) | SD* | Healthcare contact sample (N=922) | Inpatient sample (N=772) | SD* | Healthcare contact sample (N=504,682) | Inpatient sample (N=276,394) | SD* |
| Age (years) | 76 (65, 85) | 77 (66, 85) | 0.04 | 54 (36, 69) | 57 (38, 73) | 0.15 | 76 (64, 85) | 77 (65, 86) | 0.07 | 53 (36, 69) | 56 (38, 72) | 0.14 |
| Missing | 0 (0) | 0 (0) | | 0 (0) | 0 (0) | | 0 (0) | 0 (0) | | 0 (0) | 0 (0) | |
| Sex | | | | | | | | | | | | |
| Male | 427 (52) | 364 (54) | | 217,972 (44) | 113,682 (44) | | 474 (51) | 404 (52) | 0.02 | 226,381 (45) | 121,354 (44) | 0.02 |
| Female | 391 (48) | 313 (46) | 0.03 | 275,704 (56) | 147,295 (56) | 0.01 | 448 (49) | 368 (48) | | 278,301 (55) | 155,040 (56) | |
| Missing | 0 (0) | 0 (0) | | 0 (0) | 0 (0) | | 0 (0) | 0 (0) | | 0 (0) | 0 (0) | |
| Ethnicity | | | | | | | | | | | | |
| White | 652 (80) | 568 (84) | | 321,323 (65) | 188,077 (72) | | 763 (83) | 661 (86) | 0.02 | 342,474 (68) | 205,222 (74) | 0.02 |
| Non-white | 29 (4) | 24 (4) | 0.01 | 33,447 (7) | 18,246 (7) | 0.02 | 35 (4) | 28 (4) | | 31,581 (6) | 17,690 (6) | |
| Missing | 137 (17) | 85 (13) | | 138,906 (28) | 54,654 (21) | | 124 (13) | 83 (11) | 0.02 | 130,627 (26) | 53,482 (19) | 0.02 |
| Deprivation percentile | 74 (57, 89) | 74 (57, 85) | 0.01 | 74 (54, 89) | 74 (54, 89) | -0.01 | 74 (54, 88) | 73 (54, 88) | -0.02 | 74 (54, 89) | 74 (54, 89) | -0.01 |
| Missing | 2 (0) | 1 (0) | | 3,085 (1) | 1,245 (0) | | 4 (0) | 3 (0) | -0.02 | 4,065 (1) | 1,806 (1) | -0.01 |
| Rural/urban classification | | | | | | | | | | | | |
| Urban area | 535 (65) | 442 (65) | | 324,377 (66) | 171,884 (66) | | 619 (67) | 522 (68) | 0.02 | 330,264 (65) | 182,509 (66) | 0.01 |
| Rural town | 131 (16) | 112 (17) | | 78,517 (16) | 42,150 (16) | | 141 (15) | 113 (15) | | 79,908 (16) | 43,979 (16) | |
| Rural village | 150 (18) | 122 (18) | 0.02 | 87,686 (18) | 45,693 (18) | 0.01 | 158 (17) | 134 (17) | | 90,440 (18) | 48,098 (17) | |
| Missing | 2 (0) | 1 (0) | | 3,096 (1) | 1,250 (0) | | 4 (0) | 3 (0) | 0.02 | 4,070 (1) | 1,808 (1) | 0.01 |
| Catchment percentage | 94 (85, 96) | 94 (88, 96) | 0.09 | 93 (40, 96) | 93 (64, 96) | 0.07 | 95 (90, 96) | 95 (90, 96) | 0.07 | 93 (57, 96) | 93 (64, 96) | 0.03 |
| Missing | 2 (0) | 1 (0) | | 3,085 (1) | 1,245 (0) | | 4 (0) | 3 (0) | 0.07 | 4,065 (1) | 1,806 (1) | 0.03 |

**SD = Standardised Difference*

### Supplementary Figures

**Figure S1: Flowchart of the case population.**

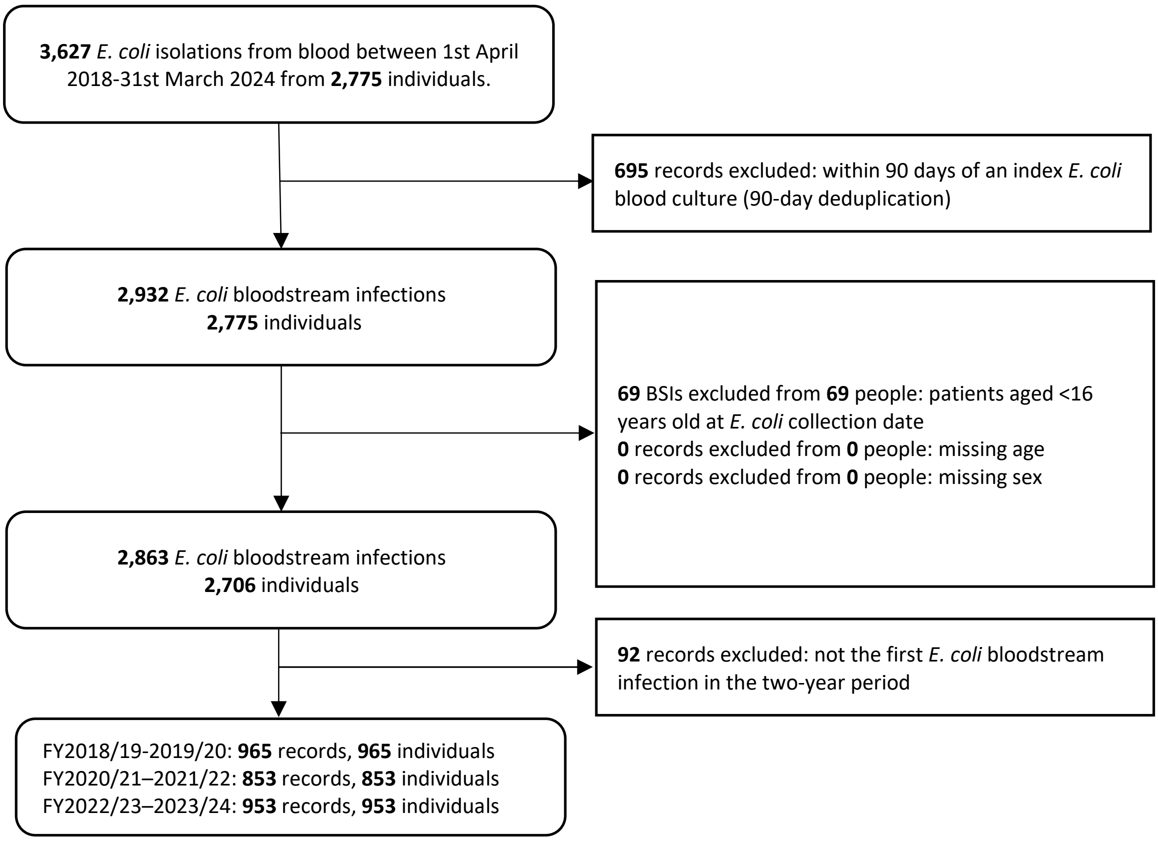

**Figure S2: Flowchart of the potential control group population.**

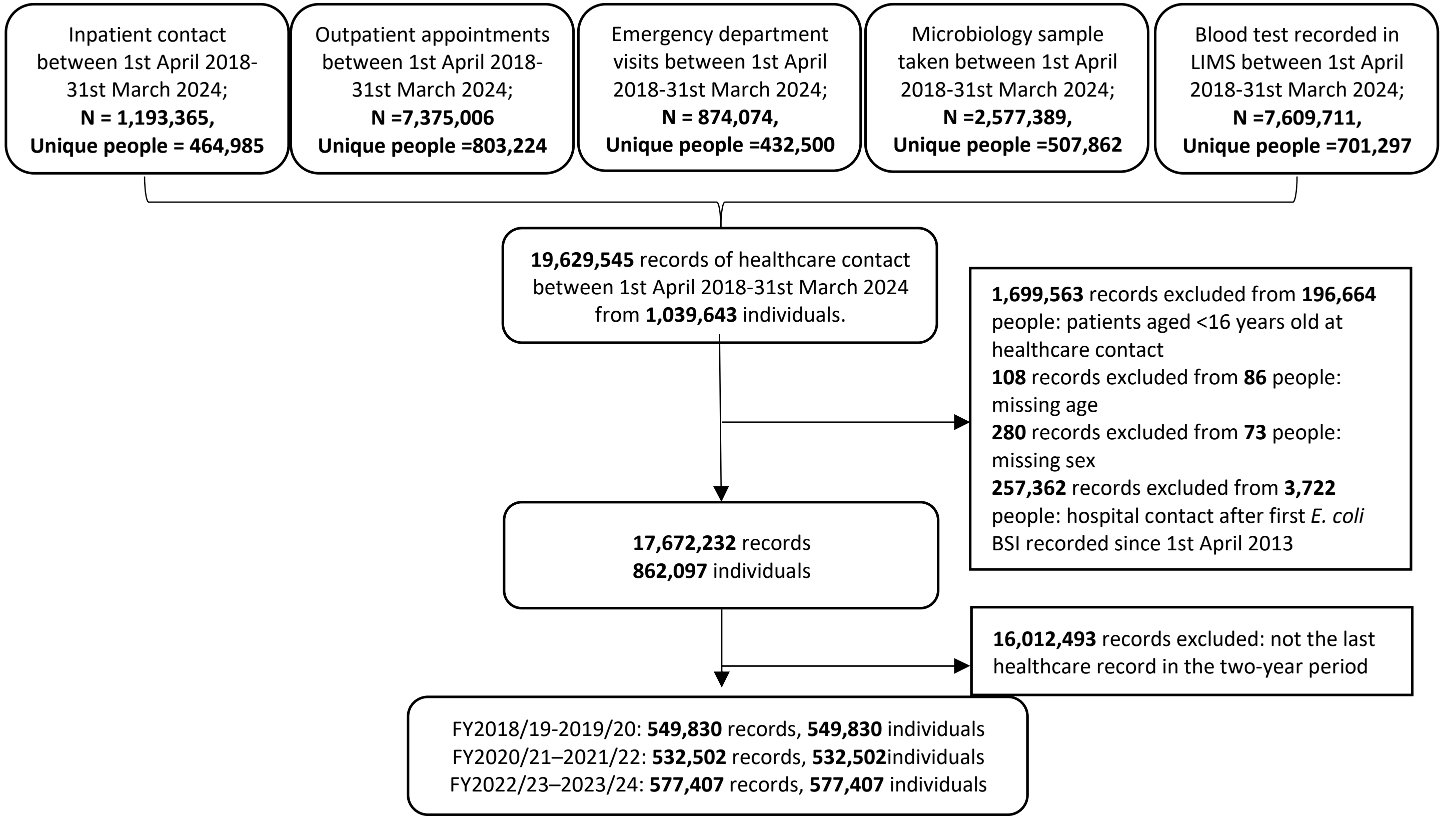

**Figure S3: A simplified Directed Acyclic Graph (DAG) illustrating potential collider bias.**

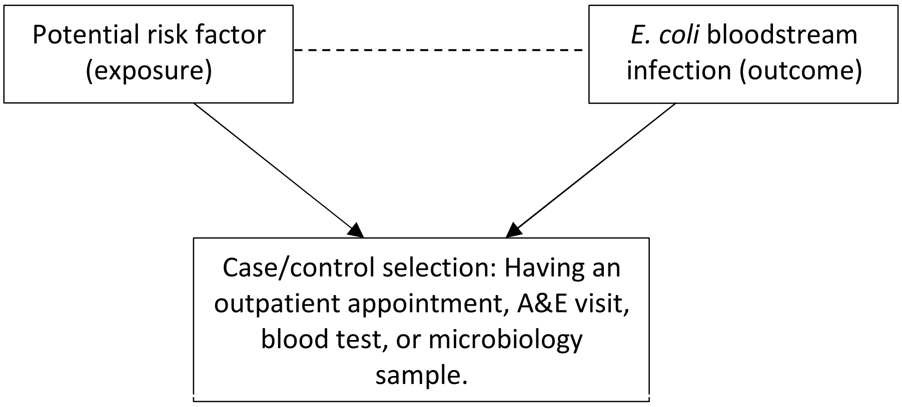

Note: *Directed arrows indicate causal effects and dotted lines indicate induced associations. Other variables and confounders are excluded for illustrative purposes only.*

**Figure S4: Catchment percentages to Oxford University Hospitals NHS Foundation Trusts for all Middle Super Output Areas as reported by the Office for Health Improvement and Disparities.**

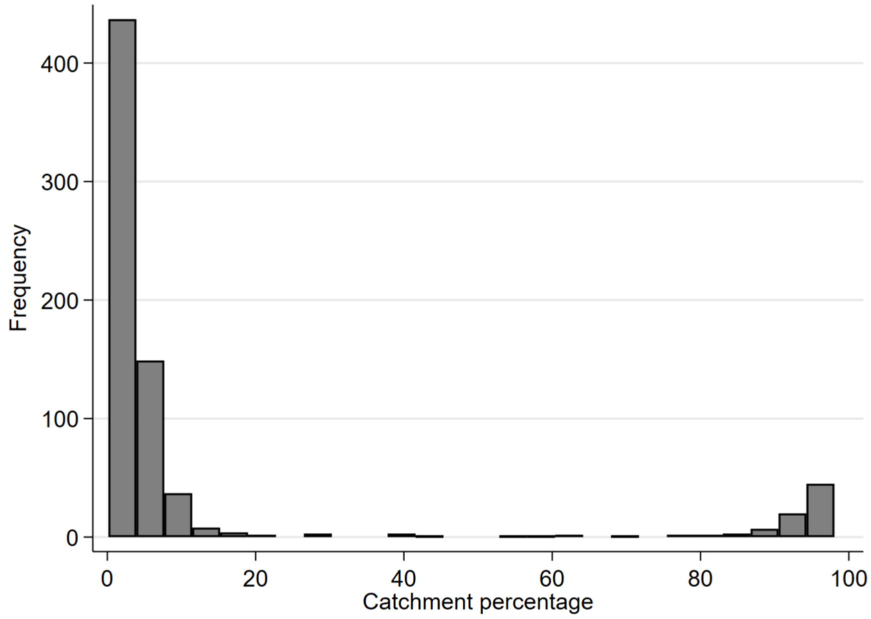

*Note: Data from https://app.box.com/s/qh8gzpzeo1firv1ezfxx2e6c4tqtrudl*

**Figure S5: Comparison of the number of diabetes cases captured in IORD versus those captured in the National Diabetes Audit for the 62 primary care practices with >80% catchment to Oxford University Hospitals NHS Foundation Trusts.**

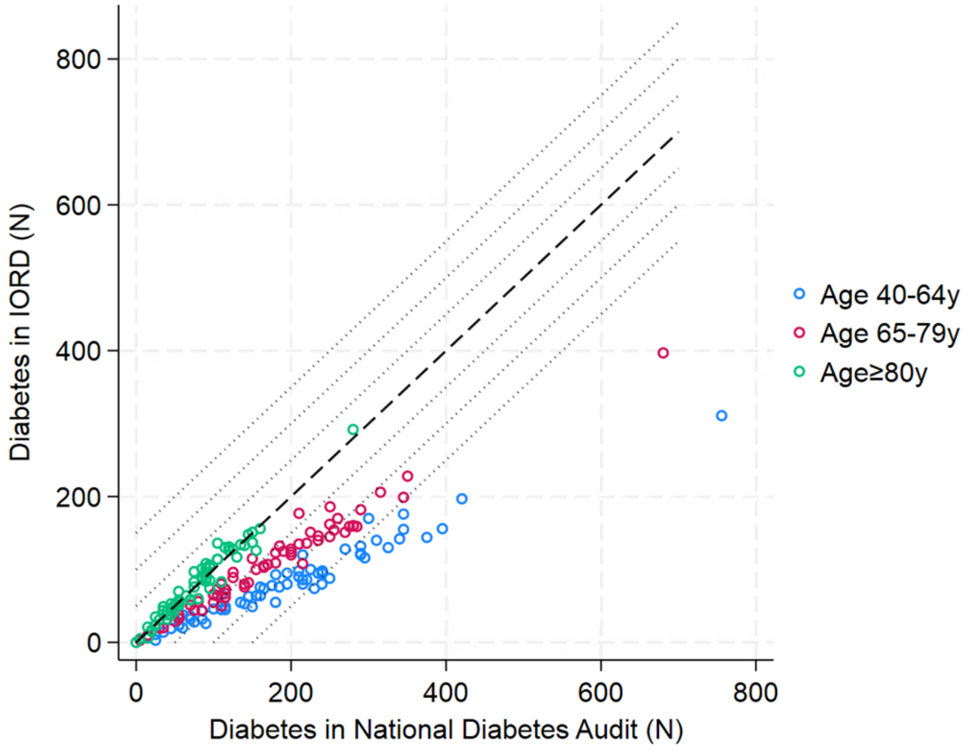

*Note: The dashed line shows agreement (y=x). The dotted lines show ±50, ±100, and ±150 diabetes cases.*

**Figure S6: Comparison of the number of people captured in IORD versus those registered at primary care for the 62 primary care practices with >80% catchment to Oxford University Hospitals NHS Foundation Trusts.**

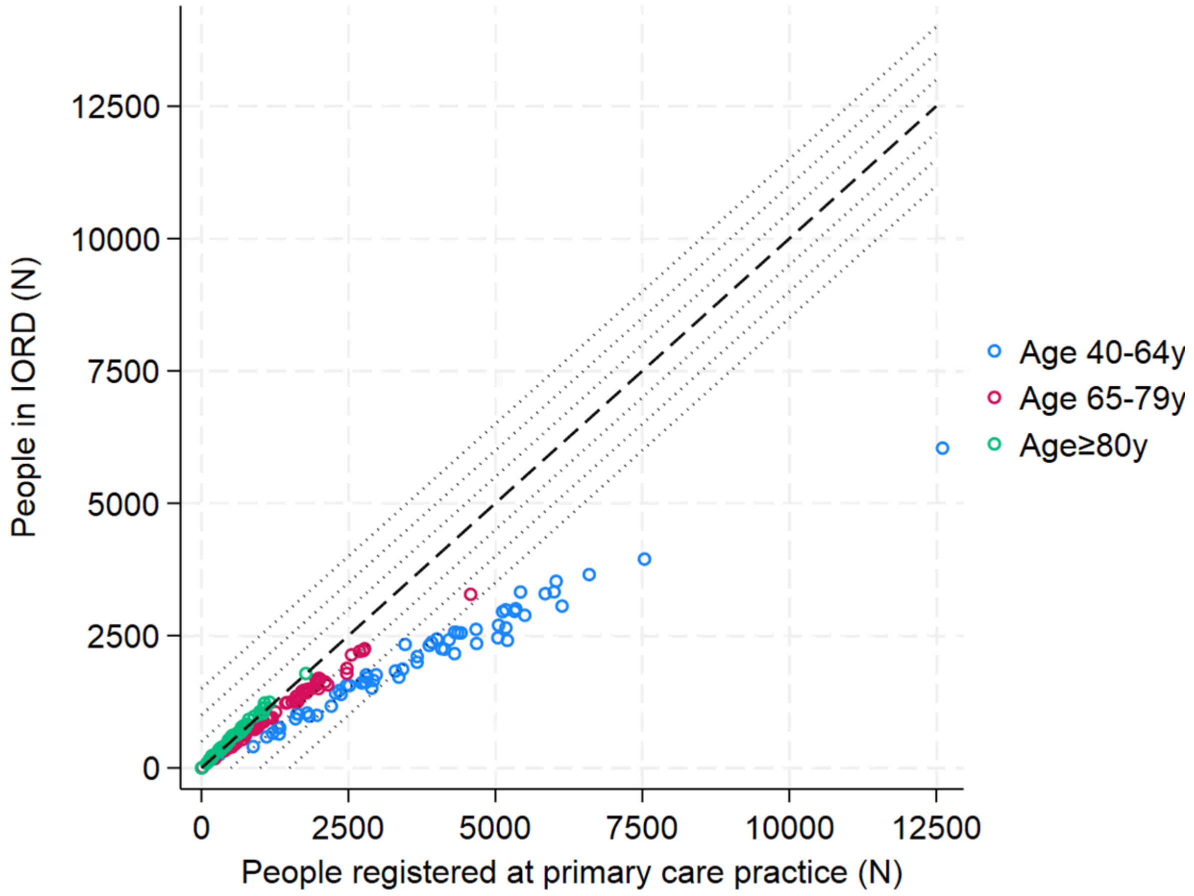

*Note: Dashed line shows agreement (y=x). Dotted lines show ±500, ±1000, and ±1500 people.*

**Figure S7: Health contact history in the previous five financial years split by the presence of *E. coli* BSI in FY2020/21-2021/22 (A) and FY2018/19-2019/20 (B).**

**A: FY2020/21-2021/22**

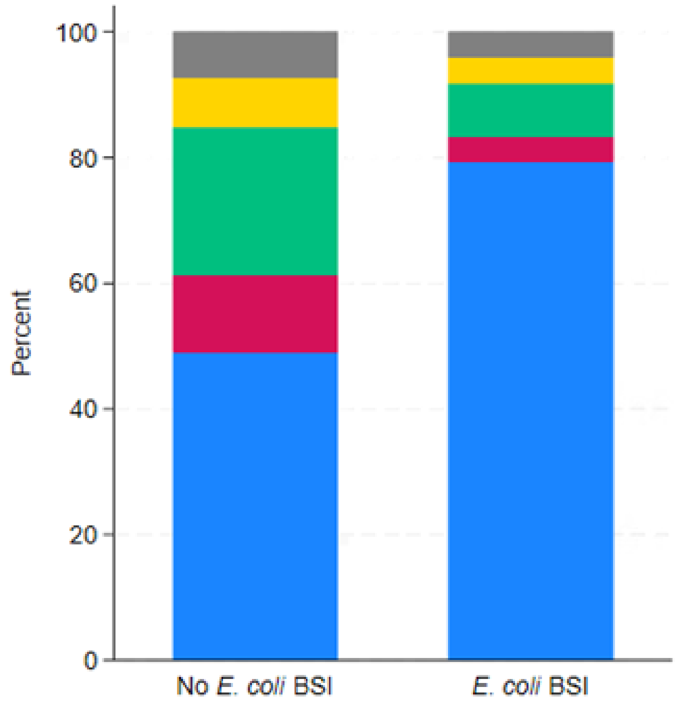

**B: FYs 2018/19-2019/20**

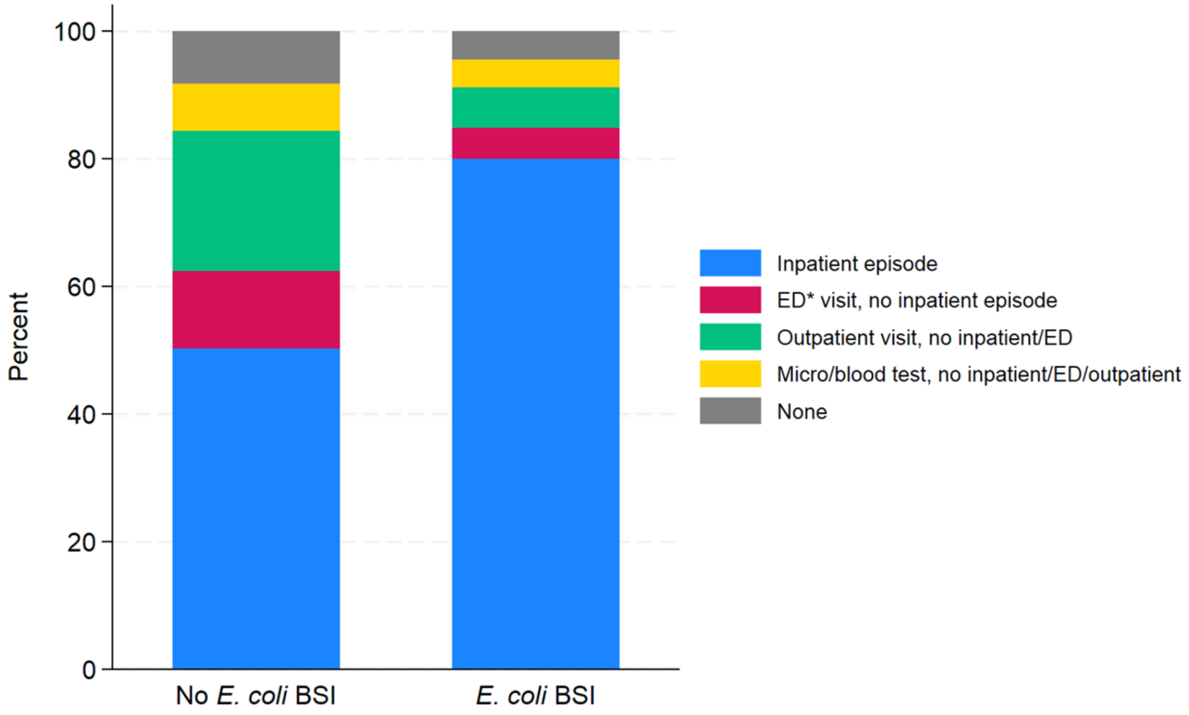

**Figure S8: Percentage of missing data for potential risk factors (left), split by hospital-linked versus inpatient samples and the presence of E. coli bloodstream infection, and standardised difference (right) between the proportion missing in the inpatient and healthcare contact samples separately for cases and controls for FY2020/21-2021/22 (A) and FY2018/19-2019/20 (B).**

**A: FY2020/21-2021/22**

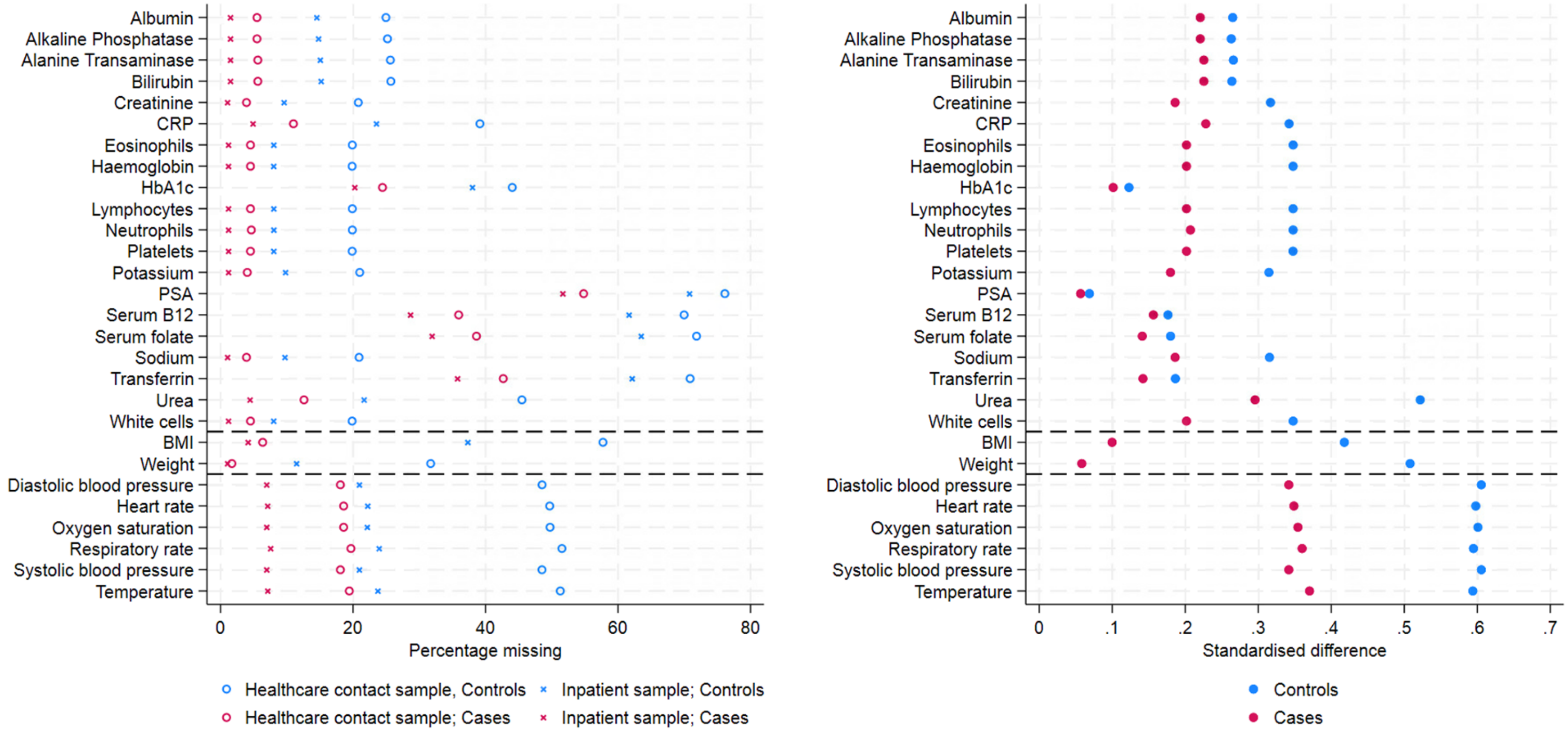

**B: FY2018/19-2019/20**

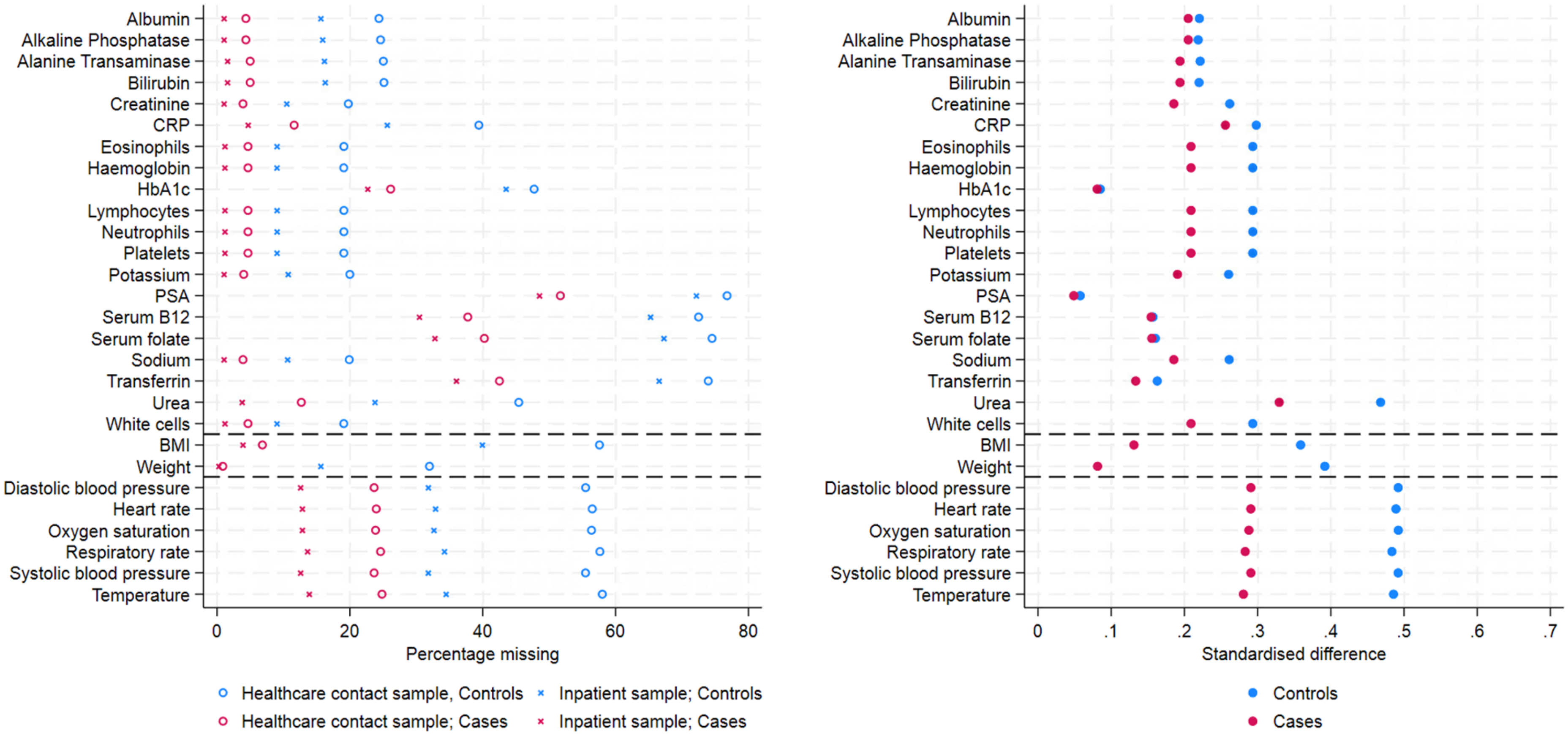

**Figure S9: Incidence rate ratios with 95% confidence intervals for core variables from Poisson models run on the hospital-linked sample (blue) and the inpatient sample (red) separately for FY2020/21-2021/22 (A) and FY2018/19-2019/20 (B).**

**A: FY2020/21-2021/22**

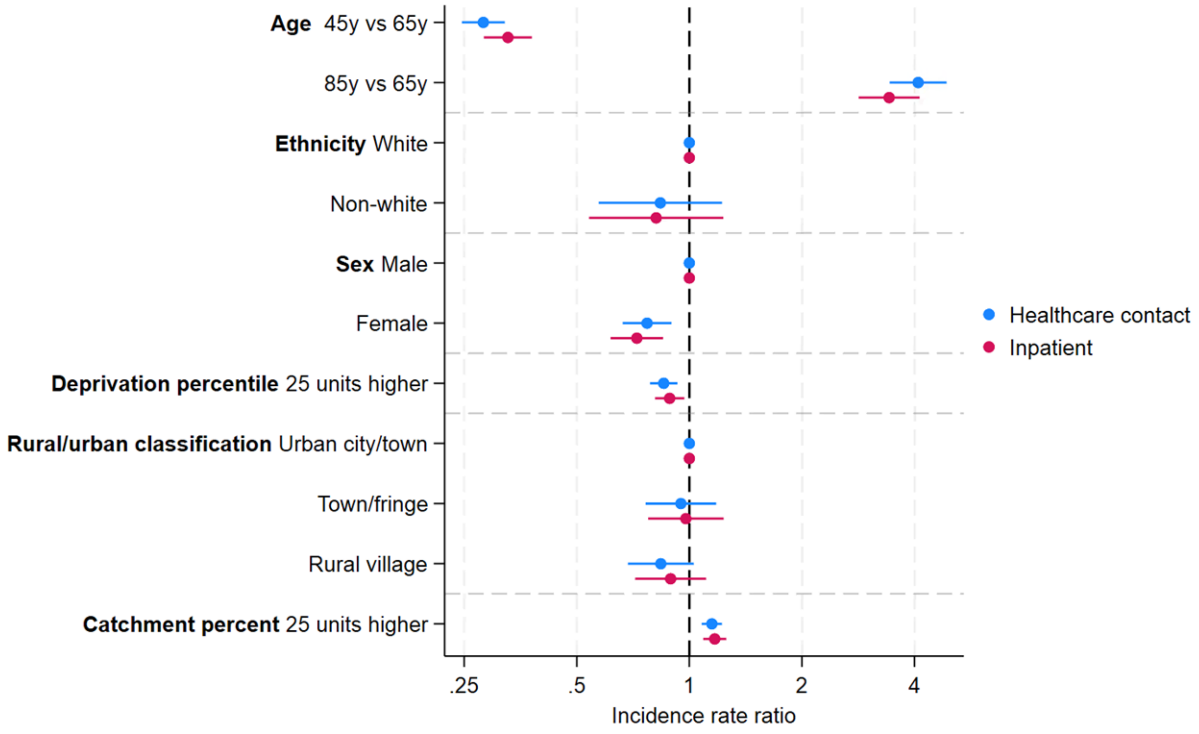

**B: FY2018/19-2019/20**

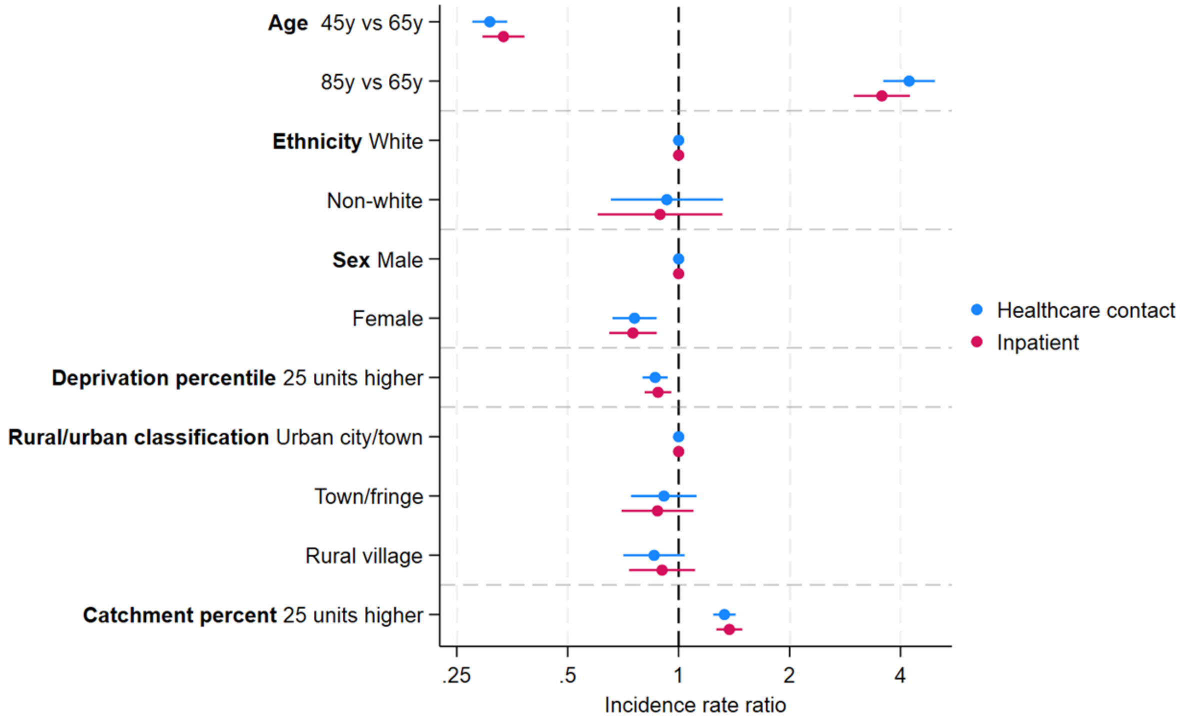

**Figure S10: Percentage of missing data for core variables and potential risk factors in *E. coli* BSI cases and controls in FY2020/21-2021/22 (A) and FY2018/19-2019/20 (B) with varying lookback.**

**A: FY2020/21-2021/22**

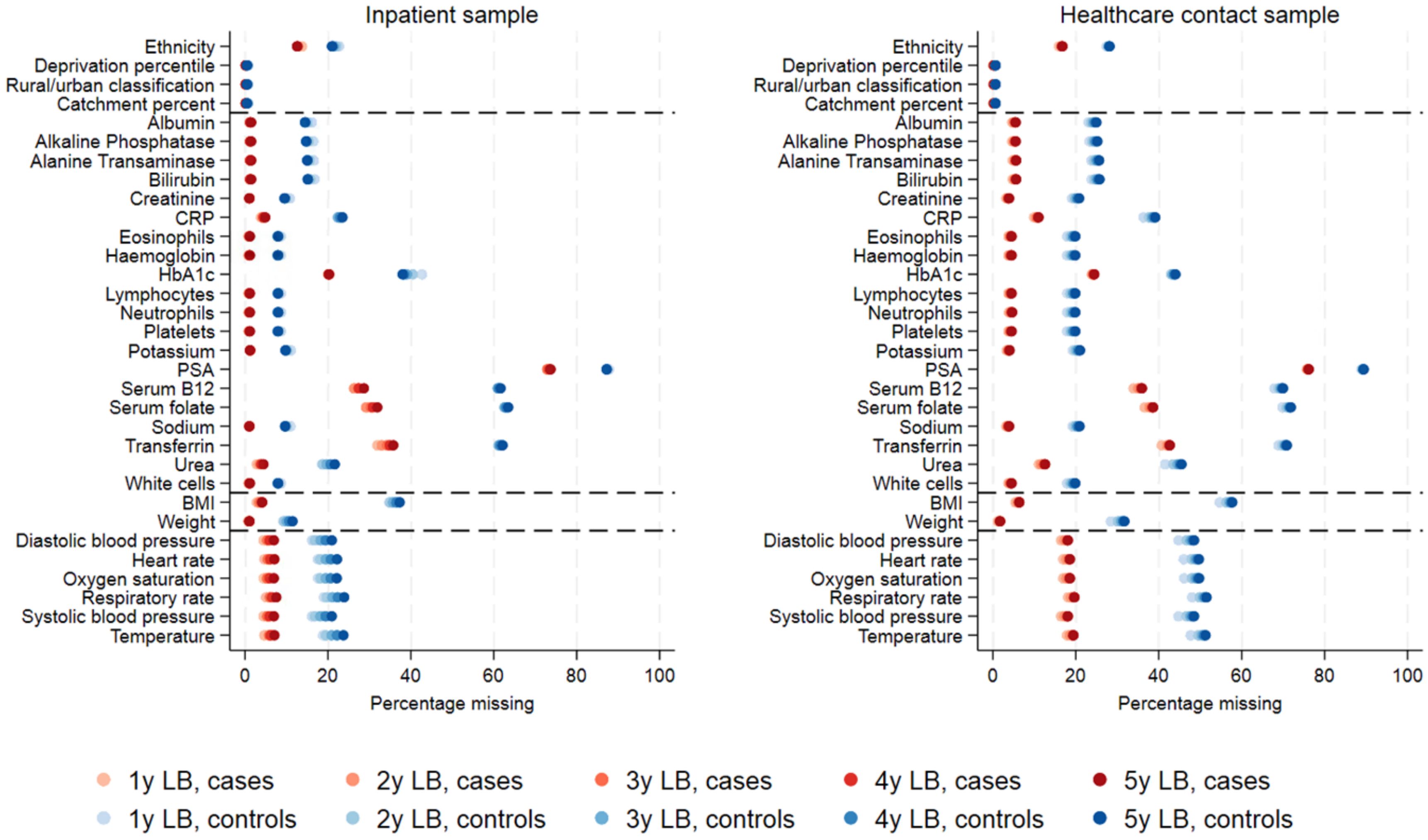

**B: FY2018/19-2019/20**

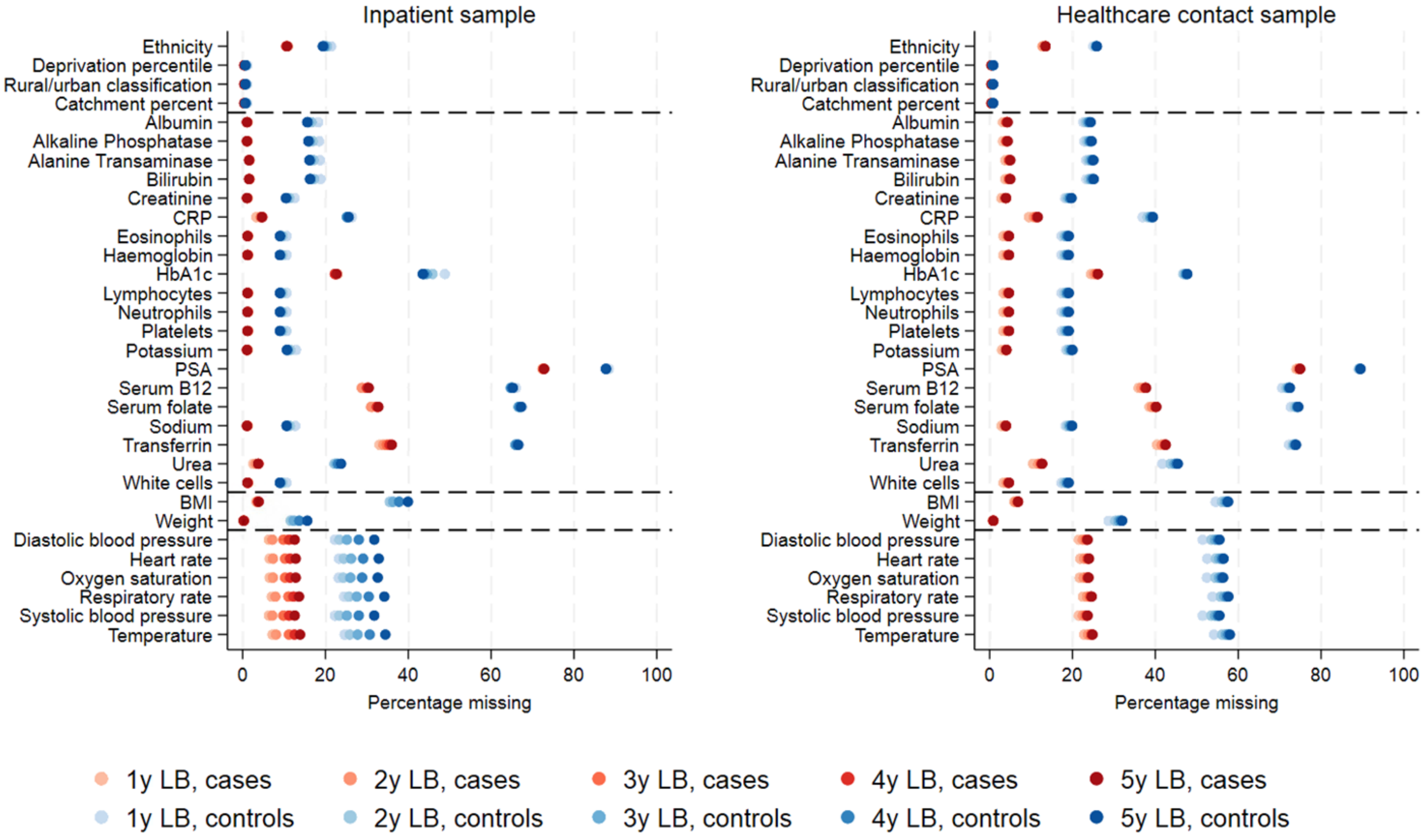

**Figure S11: Model estimates comparing different lookback lengths in the inpatient (left) and healthcare contact (right) samples in FY2020/2021-2021/22 (A) and FY2018/19-2019/20 (B).**

**A: FY2020/21-2021/22**

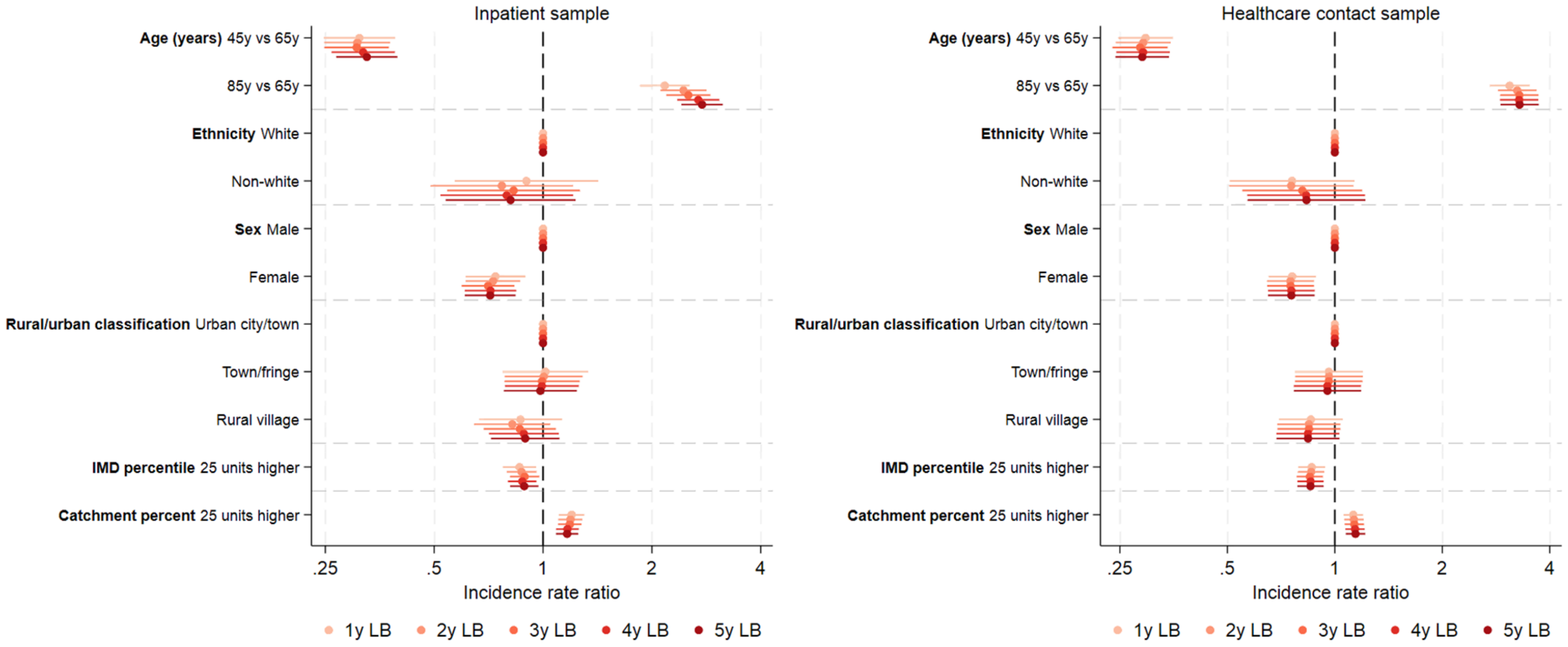

**B: FY2018/19-2019/20**

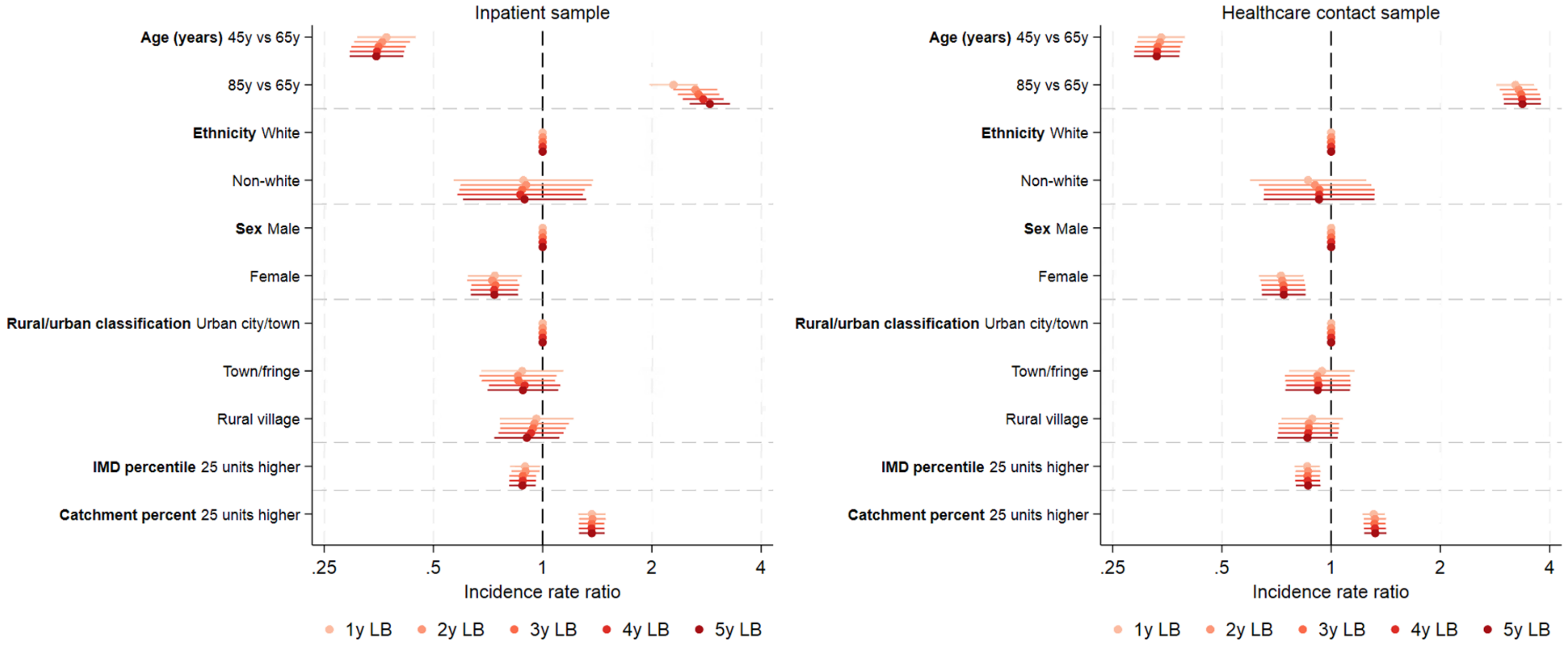
